# A Parkinson’s disease genetic risk score associates with blood DNAm on chromosome 17

**DOI:** 10.1101/2023.07.21.23293014

**Authors:** William Casazza, Samantha L. Schaffner, Fanny Artaud, Cloé Domenighetti, Laura Baglietto, Julia M. Schulze-Hentrich, Suzanne Lesage, Alexis Brice, Jean-Christophe Corvol, Sara Mostafavi, Michael S. Kobor, Alexis Elbaz, Jessica K. Dennis, DIG-PD Study Group

## Abstract

Although Parkinson’s disease (PD) coincides with altered immune functioning, there are few reproducible associations between blood DNA methylation (DNAm) and PD case-control status. Integrative analyses of genotype and blood DNAm can address this gap and can help us characterize the biological function of PD genetic risk loci. First, we tested for associations between a PD genetic risk score (GRS) and DNAm. Our GRS included 36 independent genome-wide significant variants from the largest GWAS of PD to date. Our discovery sample was TERRE, consisting of French agricultural workers (71 PD cases and 147 controls). The GRS associated with DNAm at 85 CpG sites, with 19 associations replicated in an independent sample (DIG-PD). The majority of CpG sites (73) are within a 1.5 Mb window on chromosome 17, and 36 CpG sites annotate to *MAPT* and *KANSL1*, neighboring genes that affect neurodegeneration. All associations were invariant to non-genetic factors, including exposure to commercial-grade pesticides, and omitting chromosome 17 variants from the GRS had little effect on association. Second, we compared our findings to the relationship between individual PD risk loci and blood DNAm using blood mQTL from a large independent meta-analysis (GoDMC). We found 79 CpG sites that colocalized with PD loci, and via summary Mendelian randomization analysis, we show 25/79 CpG sites where DNAm causally affects PD risk. The nine largest causal effects are within chromosome 17, including an effect within *MAPT*. Thus, all integrative analyses prioritized DNAm on chromosome 17, drawing from multiple independent data sets, meriting further study of this region.

## 1 Introduction

Parkinson’s disease (PD) is a neurodegenerative condition characterized by both motor and cognitive deficits.^1^ PD involves the aggregation of *α*-synuclein protein into structures called Lewy-bodies in the central nervous system, and ultimately results in the loss of between 60-70% of dopaminergic neurons before symptom onset.^1^ This co-occurs with changes in peripheral nervous system functioning, which can cause a diverse set of symptoms, including olfactory, cardiovascular, and gastrointestinal issues.^2, 3^ Although rare genetic mutations in *SNCA* (which encodes *α*-synuclein), *LRRK2*, *GBA*, *PINK1*, and *Parkin* can result in familial PD, up to 95% of PD cases are sporadic and are influenced by both common genetic and environmental factors.^4^

An increasing amount of evidence indicates that immune cells in blood mediate inflammatory activity underlying these symptoms, though our knowledge of which molecular changes lead to this process is incomplete.^5–7^ The relatively inexpensive array technology required for measuring DNA methylation at cytosine-guanine pairs (CpG sites) make it a hopeful target for discovering new signatures of PD in immune cells. However, population-scale epigenome-wide association studies (EWAS) of PD cases and controls report null or inconsistent results,^8–11^ with the largest PD EWAS in whole blood thus far uncovering two CpG sites associated with PD case-control status, neither of which replicated in an independent sample.^12^

Uncontrolled confounding by environmental factors and cohort-specific demographics are a possible explanation for the low replicability of PD EWAS. The environment may explain up to 81% of inter-individual variation in whole blood DNAm,^13–17^ and 73% of inter-individual variation in PD risk.^18^ Specific environmental factors that are strongly associated with PD risk include tobacco smoking and moderate alcohol consumption, which reduce the risk of PD, and head injuries and exposure to certain pesticides, which increase the risk of PD.^13, 19–21^ Sex and age are also associated with PD risk: PD is 1.4 times more prevalent in males than in females, and PD risk increases with age.^22^

Unlike the effect of PD status on DNAm, the contribution of genotype to DNAm and to PD risk, is more likely to be independent of environmental, sex, and age related variation. Genotype explains 19% of the variability in blood DNAm across the genome independent of environmental factors shared between subjects, although this varies at individual CpG sites.^16, 23^ Genotype also explains 27% of the variability in total PD risk, of which 81% (22/27) is explained by common genetic variants (i.e., variants with a minor allele frequency in Europeans of *>* 0.05) included in GWAS.^18, 24, 25^ The 90 loci that are most strongly associated with PD risk in large-scale GWAS (i.e., those with p-values <5e-8) collectively explain between 6-8% of liability to PD in independent samples.^25–27^ Thus, characterizing the function of these top loci could substantially advance our understanding of genetic risk of PD.

The objectives of this study were to: (i) measure the effects of a PD genetic risk score (GRS) on DNAm using an EWAS design; and (ii) assess the causal effect of DNAm on PD risk using summary Mendelian randomization. In our first analysis, we develop a GRS for PD using genome-wide significant variants from the existing PD GWAS, and we test its association with genome-wide DNAm in two independent PD case-control studies using an EWAS design. We test the robustness of identified associations to sex, age, smoking, alcohol consumption, prior head injury, and detailed measures of occupational pesticide exposure. In our second analysis, we identified putative causal associations between DNAm and PD risk by leveraging the existing PD GWAS, as well as a database of whole blood methylation quantitative trait loci (mQTL), which contained measures of SNP effects on DNAm identified in 32,851 participants across 36 studies.^23^ Together, these analyses identified several genomic regions where genetic risk for PD is associated with whole blood DNAm, providing multiple candidates for follow-up analyses, and a framework to complement conventional EWAS strategies.

## 2 Materials and methods

### 2.1 Study samples

We performed all discovery analysis in the TERRE case-control study. PD subjects were selected from all patients at 18 to 75 years of age who applied for national health coverage in France for PD between February 1998 to August 1999 and were affiliated in the Mutualité Sociale Agricole (MSA), an organization responsible for the reimbursement of health-related expenses for workers in the agricultural sector. Patients who previously requested health coverage for dementia or were bedridden were not included.^20, 28^ At the time of recruitment, recruiters selected up to three controls per patient from individuals enrolled in the MSA who applied for healthcare reimbursements between February 1998 and 2000, had no present diagnosis of PD, and matched for age, sex, and region of residence. Of a total of 247 PD patients and 676 controls from the initial TERRE case-control study, we accessed 88 early-stage PD patients as part of an ongoing analysis of early-stage PD (a duration *≤* 1.5 years since diagnosis) and 177 controls matched on age, sex, and region of residence for whom both genotyping and DNAm data were available (42 female PD cases, 83 female controls; 46 male PD cases, 94 male controls, Table 1).

**Table 1:**
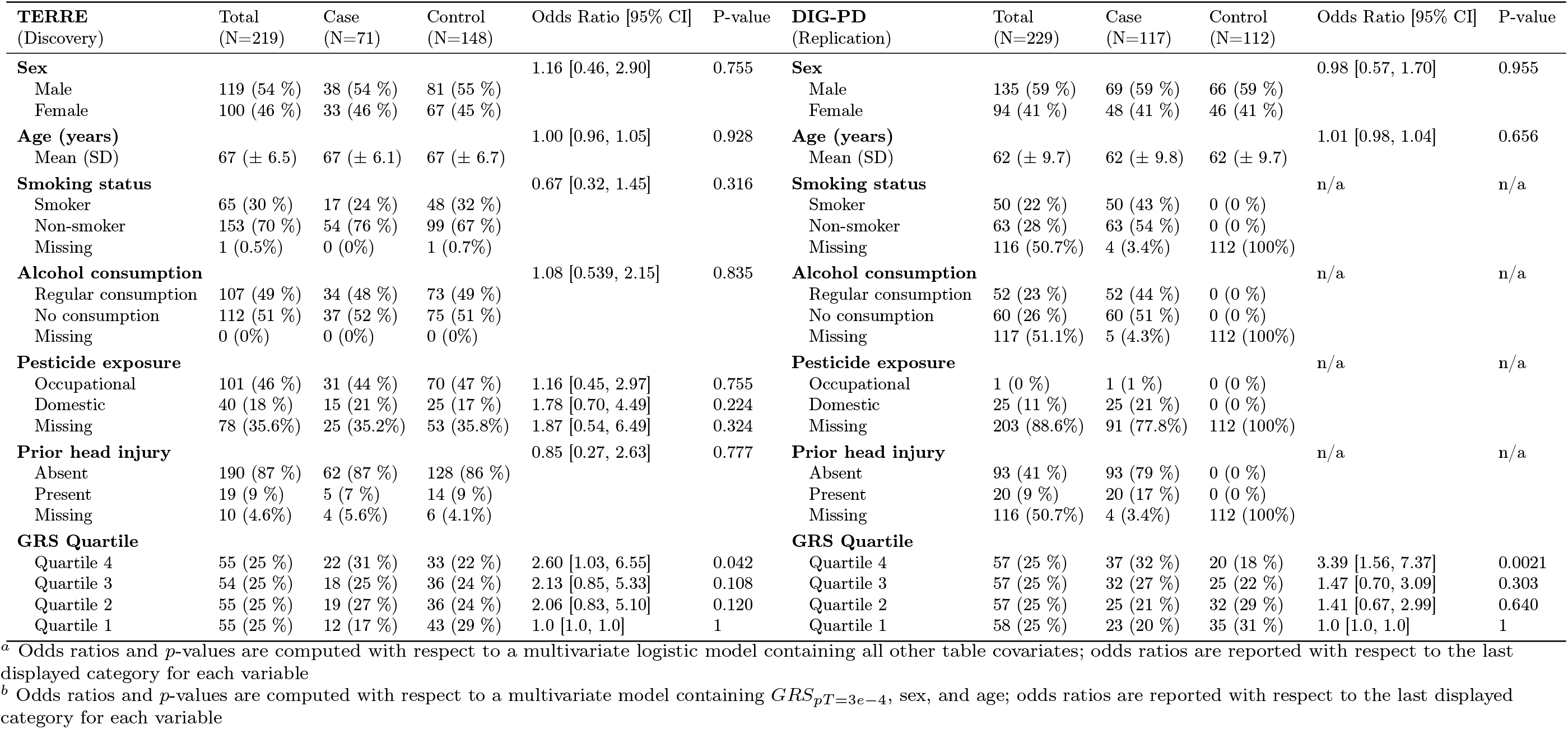
Characteristics of TERRE^*a*^ and DIG-PD^*b*^ subjects.

| TERRE<br>(Discovery) | Total<br>(N=219) | Case<br>(N=71) | Control<br>(N=148) | Odds Ratio [95% CI] | P-value | DIG-PD<br>(Replication) | Total<br>(N=229) | Case<br>(N=117) | Control<br>(N=112) | Odds Ratio [95% CI] | P-value |
| --- | --- | --- | --- | --- | --- | --- | --- | --- | --- | --- | --- |
| **Sex** |  |  |  | 1.16 [0.46, 2.90] | 0.755 | **Sex** |  |  |  | 0.98 [0.57, 1.70] | 0.955 |
| Male | 119 (54 %) | 38 (54 %) | 81 (55 %) |  |  | Male | 135 (59 %) | 69 (59 %) | 66 (59 %) |  |  |
| Female | 100 (46 %) | 33 (46 %) | 67 (45 %) |  |  | Female | 94 (41 %) | 48 (41 %) | 46 (41 %) |  |  |
| **Age (years)** |  |  |  | 1.00 [0.96, 1.05] | 0.928 | **Age (years)** |  |  |  | 1.01 [0.98, 1.04] | 0.656 |
| Mean (SD) | 67 (± 6.5) | 67 (± 6.1) | 67 (± 6.7) |  |  | Mean (SD) | 62 (± 9.7) | 62 (± 9.8) | 62 (± 9.7) |  |  |
| **Smoking status** |  |  |  | 0.67 [0.32, 1.45] | 0.316 | **Smoking status** |  |  |  | n/a | n/a |
| Smoker | 65 (30 %) | 17 (24 %) | 48 (32 %) |  |  | Smoker | 50 (22 %) | 50 (43 %) | 0 (0 %) |  |  |
| Non-smoker | 153 (70 %) | 54 (76 %) | 99 (67 %) |  |  | Non-smoker | 63 (28 %) | 63 (54 %) | 0 (0 %) |  |  |
| Missing | 1 (0.5%) | 0 (0%) | 1 (0.7%) |  |  | Missing | 116 (50.7%) | 4 (3.4%) | 112 (100%) |  |  |
| **Alcohol consumption** |  |  |  | 1.08 [0.539, 2.15] | 0.835 | **Alcohol consumption** |  |  |  | n/a | n/a |
| Regular consumption | 107 (49 %) | 34 (48 %) | 73 (49 %) |  |  | Regular consumption | 52 (23 %) | 52 (44 %) | 0 (0 %) |  |  |
| No consumption | 112 (51 %) | 37 (52 %) | 75 (51 %) |  |  | No consumption | 60 (26 %) | 60 (51 %) | 0 (0 %) |  |  |
| Missing | 0 (0%) | 0 (0%) | 0 (0%) |  |  | Missing | 117 (51.1%) | 5 (4.3%) | 112 (100%) |  |  |
| **Pesticide exposure** |  |  |  |  |  | **Pesticide exposure** |  |  |  | n/a | n/a |
| Occupational | 101 (46 %) | 31 (44 %) | 70 (47 %) | 1.16 [0.45, 2.97] | 0.755 | Occupational | 1 (0 %) | 1 (1 %) | 0 (0 %) |  |  |
| Domestic | 40 (18 %) | 15 (21 %) | 25 (17 %) | 1.78 [0.70, 4.49] | 0.224 | Domestic | 25 (11 %) | 25 (21 %) | 0 (0 %) |  |  |
| Missing | 78 (35.6%) | 25 (35.2%) | 53 (35.8%) | 1.87 [0.54, 6.49] | 0.324 | Missing | 203 (88.6%) | 91 (77.8%) | 112 (100%) |  |  |
| **Prior head injury** |  |  |  | 0.85 [0.27, 2.63] | 0.777 | **Prior head injury** |  |  |  | n/a | n/a |
| Absent | 190 (87 %) | 62 (87 %) | 128 (86 %) |  |  | Absent | 93 (41 %) | 93 (79 %) | 0 (0 %) |  |  |
| Present | 19 (9 %) | 5 (7 %) | 14 (9 %) |  |  | Present | 20 (9 %) | 20 (17 %) | 0 (0 %) |  |  |
| Missing | 10 (4.6%) | 4 (5.6%) | 6 (4.1%) |  |  | Missing | 116 (50.7%) | 4 (3.4%) | 112 (100%) |  |  |
| **GRS Quartile** |  |  |  |  |  | **GRS Quartile** |  |  |  |  |  |
| Quartile 4 | 55 (25 %) | 22 (31 %) | 33 (22 %) | 2.60 [1.03, 6.55] | 0.042 | Quartile 4 | 57 (25 %) | 37 (32 %) | 20 (18 %) | 3.39 [1.56, 7.37] | 0.0021 |
| Quartile 3 | 54 (25 %) | 18 (25 %) | 36 (24 %) | 2.13 [0.85, 5.33] | 0.108 | Quartile 3 | 57 (25 %) | 32 (27 %) | 25 (22 %) | 1.47 [0.70, 3.09] | 0.303 |
| Quartile 2 | 55 (25 %) | 19 (27 %) | 36 (24 %) | 2.06 [0.83, 5.10] | 0.120 | Quartile 2 | 57 (25 %) | 25 (21 %) | 32 (29 %) | 1.41 [0.67, 2.99] | 0.640 |
| Quartile 1 | 55 (25 %) | 12 (17 %) | 43 (29 %) | 1.0 [1.0, 1.0] | 1 | Quartile 1 | 58 (25 %) | 23 (20 %) | 35 (31 %) | 1.0 [1.0, 1.0] | 1 |

<sup>a</sup> Odds ratios and $p$ -values are computed with respect to a multivariate logistic model containing all other table covariates; odds ratios are reported with respect to the last displayed category for each variable
<sup>b</sup> Odds ratios and $p$ -values are computed with respect to a multivariate model containing $GRS_{pT=3e-4}$ , sex, and age; odds ratios are reported with respect to the last displayed category for each variable

A neurologist specializing in movement disorders diagnosed PD in subjects according to the UK Parkinson’s Disease Society Brain Bank criteria, and was defined as the presence of parkinsonism with exclusion of drug-induced phenotypes or further nervous system involvement.^29^ No PD patients in the TERRE study had known familial risk factors in *GBA* (E326K) or *LRRK2* (G2019S). Likewise, a physician interviewed each participant to record demographic data and administer the Mini Mental State Examination (MMSE).^30^ Recruiters initially assessed occupational pesticide exposure using a questionnaire designed for this study. For those that responded yes, this was followed by conducted occupational health interviews to obtain detailed information on patients’ history of pesticide exposure, including the number and type of farms where the individuals had worked, which pesticides they had personally sprayed, the frequency, duration, and method of spraying, and number of years that they were exposed.^20, 31, 32^ We imputed missing values for individuals exposed to pesticides as previously described.^20^.

We assessed replication the drug interaction with genes in Parkinson’s Disease (DIG-PD) case-control study.^33^ Cases (disease duration *≥* 5 years at recruitment) were recruited from 4 French university hospitals and 4 general hospitals between May 2009 and July 2013. Eligible patients had previously been diagnosed with PD by movement disorder neurologists, according to the UK Parkinson’s Disease Society Brain Bank criteria.^26, 34^ Patients were followed up annually for 7 years, but for the purpose of this study we only used data demographic data collected from physicians at recruitment. We excluded individuals with familial PD mutations determined from genotyping data: five individuals with the E326K risk factor in *GBA* and two individuals with a mutation in *LRRK2* (G2019S). From 411 cases, we selected 117 cases with both DNAm and genotyping measurements. We compared these patients to 112 age-and sex-matched controls without PD who had both DNAm and genotyping measurements, and for whom no additional phenotypic data was available.^35^

### 2.2 Genotype processing and imputation

All genotype processing steps in TERRE and DIG-PD were based on the RICOPILI pipeline.^36^ In TERRE, cases and controls were genotyped on the Illumina NeuroChip array (486,137 SNPs). We used plink 1.9^37^ for pre-imputation QC in which we removed samples with a mismatch between recorded and genotyped sex; missingness *>* 0.02 and *F_het_ >* 0.2. We also removed SNPs that were strand-ambiguous, had a call rate *<* 0.05, and a minor allele frequency (MAF) *<* 0.01

Samples and SNPs passing these pre-imputation QC steps were used to compute genotype principal components (PCs). For this analysis, we removed SNPs in high linkage disequilibrium (i.e., -indep-pairwise 200 0.2 in plink 1.9 was run twice), SNPs with an MAF *<* 0.05, and SNPs in highly recombinant regions: the HLA region, chr6:28,477,797-33,448,354 in GRCh37, and regions of long range LD.^38^ As we later restricted our analysis to SNPs with available GWAS summary statistics, we excluded the sex chromosomes in this analysis.^25, 39^ We filtered samples to ensure identity by descent did not exceed an *|*F_ST_*|>* 0.2 between samples, and computed 10 genotyping PCs using FastPCA.^40^

Before proceeding with imputation, we removed variants associated with genotype batch (Bonferroni corrected *p <* 0.05) and variants with a Hardy-Weinberg equilibrium *p <* 1 *∗* 10*^−^*^10^ using bcftools.^41^ We then imputed genotypes to the 1000 genomes phase 3 European reference panel using the Michigan imputation server,^42^ and filtered variants to have an imputation quality *R*2 *>* 0.3 and a MAF within 0.1 of the 1000 genomes phase 3 European population. At this stage we generated an additional set of genotypes for GRS analysis, in which we hard-called SNPs at a threshold of 0.1 (–hard-call-threshold 0.1 in plink2^37^). Lastly, we removed imputed SNPs associated with their genotype batch, and we checked for inflation of the association between genotype and PD by logistic regression accounting for sex, age at study enrollment, and three genotyping PCs to account for cryptic relatedness between sample (which collectively explained 15.5% of variation in genotype).^43^ We observed deflation in the association between genotype and PD status, which is likely reflective of low sample size (Figures S1A,B; with genomic inflation factor *λ_GC_* = 1.06, Figure S1C). The total number of SNPs available for analysis after quality control was 8,354,189.

DIG-PD cases were genotyped on the Illumina Infinium Multi-Ethnic Global Array (MEGA; 1,779,819 SNPs). DIG-PD controls were genotyped on the Illumina NeuroChip Array. We used the same pre-imputation QC and genotype PC analysis as in TERRE in both DIG-PD cases and controls. We took the following measures during imputation to account for cases and controls being mapped on different platforms, ultimately merging samples after all imputation steps.^44^ Genotypes were imputed separately in cases and controls using the Michigan imputation server.^45^ We first imputed SNPs to the 1000 Genomes European reference panel, and filtered resulting imputed SNPs at a stringent *R*2 *>* 0.9, followed by a second round of imputation and filtering at an *R*2 *>* 0.9. After imputation, we merged cases and controls and filtered to SNPs with a MAF within 0.1 of the 1000 genomes phase 3 European population. We then generated a set of hard-called SNPS, and then checked for an inflation of association between genotype and PD with a logistic regression accounting for sex, age at study enrollment, and three genotyping PCs (explaining 16.1% and 16.0% of variation in cases and controls respectively; Figures S2 A-D). After all quality control measure, 881,726 SNPs were available for analysis, which is less than what is available in TERRE due to the stringent quality control required to harmonize our early-stage PD cases with controls. Although this is substantially fewer, we are confident that our procedure reduces biases induced by cases and controls being genotyped on separate platforms. To assess this, we repeated our imputation procedure at less stringent R2 *>* 0.3 and R2 *>* 0.80, and assessed inflation of summary statistics (*λ_GC_* = 1.15 and *λ_GC_*= 1.11, respectively, Figures S2 E, F). At an R2 *>* 0.9, inflation was substantially lower (*λ_GC_* = 1.04; Figure S2G).

### 2.3 DNA methylation processing

For the TERRE and DIG-PD studies, participants provided a blood sample at recruitment. From these blood samples, DNAm was assessed at 853,307 CpG and 2,880 CNG (where N represents any nucleotide) sites using the Illumina HumanMethylationEPIC BeadChip array. We read raw idat files into R 3.6 for data analysis using the minfi package.^46^ We applied the following sample filtering procedures separately in each sample: quality control checks using control_metrics from the ewastools package, methylated/unmethylated intensity checks using getQC from the minfi package, clustering of SNP control probe betas, SNP-based outlier detection using snp_outliers from ewastools, removal of samples with detection *p*-value *>* 0.01 at *>* 5% of probes, removal of samples with bead count *<* 3 at *>* 1% of probes, removal of samples with average intensity below the mean intensity *−*2 *∗* standard deviation, outlier detection using detectOutlier from the lumi package and outlyx from the wateRmelon package, removal of samples with inter-sample Pearson correlation *<* 0.95, removal of samples clustering away from centroids by principal component analysis (PCA), removal of samples with extreme deviations between reported age and predicted DNAm age using the Horvath clock, and removal of samples with atypical beta value distributions.^14, 46–49^ Additionally, we confirmed reported sex using PCA on X chromosome beta values, percentage of missing values on the Y chromosome, the getSex function from the minfi package, and X/Y chromosome copy number using the conumee package.^46, 50, 51^ After applying all sample filtering procedures, we removed 34 samples from TERRE (231 remaining), and we removed 64 samples from DIG-PD (278 remaining). The datasets were later subset to individuals with matched genotyping data, and individuals without familial mutations (71 cases, 147 controls in TERRE; 117 cases, 112 controls in DIG-PD; Table 1).

To control for unwanted sources of technical variation in DNAm, we performed functional normalization in each study using adjustedFunnorm from the wateRmelon package.^52^ After normalization, we removed low-quality probes from the datasets to ensure quality control at the individual CpG level. In each study, we removed probes with bead count *≤* 3 and *>* 5% missing values; probes with detection *p*-values *≥* 0.01; cross-hybridizing and polymorphic probes; and SNP control probes.^49^ This left 803,777 probes in TERRE, 803,734 probes in DIG-PD. We retained X and Y chromosome probes to allow for possible analysis on sex-chromosomes.

To assess and account for the effects of differing blood (immune) cell proportions on DNAm levels in each study, we predicted cell type proportions in raw data using the original IDOL library including six immune cell types.^53^ For TERRE, we noted differences (nominal two-sided Wilcoxon *p <* 0.05) in cell type proportion between cases vs. controls for NK cells and B cells. However, these differences are not significant after adjusting for multiple test correction (FDR *<* 0.05; Figure S3A). In males vs. females for CD8^+^ T cells, B cells, and monocytes (of which only the differences in B cell and monocyte proportions were significant after multiple test correction, FDR *<* 0.05; Figure S3B). For DIG-PD, we noted differences in cell type proportion between cases vs. controls for CD8^+^ T cells, NK cells, B cells, and Neurons (Figure S3C), and in males vs. females for CD4^+^ T cells, B cells, and monocytes (monocyte proportions were not significantly different at FDR *<* 0.05; Figure S3D).

In addition to these differences in cell type proportions by case/control status and sex, we also observed a correlation between batch (plate and row) and sex. In order to simplify our analyses, we opted to account for global variation in DNAm, i.e., those that influence DNAm across all CpG sites, using the top 10 principal components of DNAm in each study.^54–56^ We observed moderate to high correlation (Pearson’s *r* = 0.4 *−* 0.9) between cell type proportion estimates and the top 5 DNAm PCs for each study(Figure S4). As for batch, variables, we observed similarly high correlation between plate and the top 10 DNAm PCs for each study, but low correlation between row and top 10 DNAm PCs, suggesting that it was not a major source of global variation in DNAm in either study.

### 2.4 Calculating Parkinson’s disease genetic risk scores

We calculated GRS of Parkinson’s status using a PRSice-2.^57^ They were computed separately on both TERRE and DIG-PD using summary statistics from a publicly available GWAS meta-analysis of 33,674 PD cases and 449,056controls.^25^ We applied the following filters to these summary statistics: a minor allele frequency (MAF) *>* 0.01, removal of strand ambiguous SNPs, and removal of duplicate SNPs. LD for clumping and thresholding was estimated using the 1000 Genomes Phase 3 European reference panel. We ran PRSice-2 against hard-called SNPs (see above) at all default *p*-value thresholds(*pT*), e.g., from *pT* = 5 *∗* 10*^−^*^8^ to *pT* = 0.5 assessing all *pT* intervals of 5 *∗* 10*^−^*5, and *pT* = 1. We denote a GRS constructed from SNPs significantly associated with PD at a threshold of *K* as *GRS_pT_* _=*K*_.

As a benchmark, we first developed GRS that best predicted PD status in TERRE, as measured by the difference in *R*^2^ value from a logistic regression model (see below) and the *baseline* + *GRS_pT_* _=*K*_

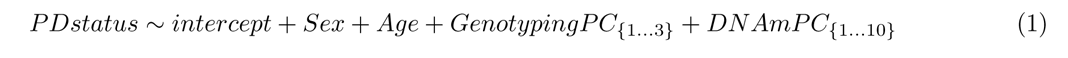

In other words, we selected SNPs such that the GRS associated with PD status independent of other covariates affecting global DNAm. We assessed the fit of each GRS in all samples (i.e., cross-sex), male samples, and female samples. GRS computed at a *pT <* 0.125 had a larger *R*^2^ in male samples compared to the cross-sex sample and GRS with *pT >* 0.125 had a larger *R*^2^ in female samples compared to the cross-sex sample(Figure S5A,B). To estimate the proportion of heritability explained on the liability scale (*h*^2^) we assume a 0.5% prevalence of PD, which is approximately the global rate of PD in people between the ages of 60-70.^58^

In several psychiatric traits, EWAS of GRS, rather than case control status, showed increased power to detect changes in DNAm. Moreover, these associations appeared to be independent of non-genetic factors which confounded EWAS of case-control status in the same conditions.^55, 59, 60^ However, in the case of PD, genetic correlation between PD risk in males vs. females (*r_g_*= 0.877, SE = 0.0699) is low compared to male-female genetic correlation across traits in the UK Biobank, which warrants additional attention to sex in genetic analyses of PD.^39, 61^ As such, we also compared sex-specific genetic risk to sex-agnostic genetic risk for PD in male vs. female samples in TERRE (Supplementary Material Section S1) using sex-stratified GWAS results.^39^ However, since the sex-stratified GWAS results included fewer individuals, they were less powerful, and we found that these GRS explained less variability in PD than those computed using cross-sex GWAS summary statistics. We therefore used GRS derived from cross-sex summary statistics in all downstream analyses.

### 2.5 Epigenome-wide association with Parkinson’s GRS

Association between GRS and DNAm M-values (i.e., logit-transformed beta values)^62, 63^ was computed using limma^64^ according to the following linear model:

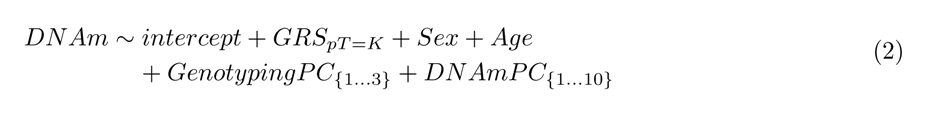

We then sought to find the pT that maximized the number of significant associations with an FDR *<* 0.05 and a difference of M-values > 1.5, as opposed to GRS that best predicted PD status. A *pT* = 5*e −* 8 was the best fit in the cross-sex sample, and in males and females separately, as computed cross-sex PD GWAS summary statistics.^25, 62^

We tested whether demographic variables and other non-genetic factors (i.e., exposures), or their interaction with the GRS, influenced the association between our cross-sex GRS and DNAm in TERRE. We assessed the association of each exposure with DNAm accounting for *GRS_pT_* _=5*e*_*_−_*_8_ and the interaction of *GRS_pT_* _=5*e*_*_−_*_8_ with the exposure (*GRS × Exposure*) using the following linear model:

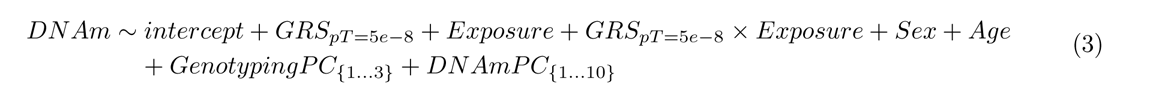

We assessed smoking status, alcohol use, age, sex, self-reported previous head injury, and 69 different occupational pesticide and heavy metal exposures surveyed in TERRE (Table 1, S1). As information was not available on all pesticide exposures for each sample, we used data imputed via SAS PROC MI,^20^ and used mice^65^ to average our results over 10 imputations. For non-imputed data, we used the limma package.^64^

### 2.6 Colocalization analysis

We performed colocalization analysis with GWAS *p*-values from extcitenallsIdentificationNovelRisk2019 and mQTL *p*-values from GoDMC, a large (N=32,851 subjects across 36 studies) mQTL meta-analysis in blood measured computed using Illumina’s 450K HumanMethylation Array data. We used coloc to compute colocalization between cis-mQTL (*p <* 5 *∗* 10*^−^*^8^) and GWAS variants for each CpG site within 1 Mb of the representative SNP of each GWAS locus.^23, 66, 67^ Next, we derived independent GWAS loci over significant SNPs (*p <* 5 *∗* 10*^−^*^8^) with plink 1.9 -clump (default parameters of 250kb and *r*^2^ *>* 0.5 in the 1000 Genomes phase3 European reference panel^37, 42^). This resulted in 79 independent loci. To ensure GoDMC mQTL were representative of associations present in TERRE, we assessed replication between mQTL computed in TERRE and matching mQTL from GoDMC. Replication was high as assessed using *π*_1_ statistic (the proportion of significant associations in TERRE with non-null effects in GoDMC *π*_1_ = 0.95, Supplementary Material Section S2). We called colocalization at a posterior probability *P* (*H*_4_) *>* 0.9, which corresponds to the probability of the same SNP associating with both PD risk and a DNAm.

### 2.7 Summary Mendelian Randomization

In order to test whether colocalized mQTL showed evidence of a causal association between levels of DNAm and PD, we ran a summary Mendelian randomization (SMR) analysis.^68, 69^ Briefly, SMR estimates the association between an DNAm and PD risk ^^^*b_xy_* from the effect of genotype on DNAm, *β*^^^*_zx_* and the effect of genotype on PD risk ^^^*b_zy_* as

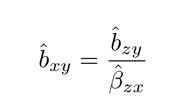

We computed this estimate using the top mQTL for a given CpG site and its corresponding association in GWAS. In order to estimate significance of this estimate, it makes use of the Delta method to compute the variance of ^^^*b_xy_*, and assumes that *β*^^^*_zx_* and ^^^*b_zy_* are estimated from independent samples. Again, we made use of mQTL computed in GoDMC^23^ and GWAS summary statistics from extcitenallsIdentificationNovelRisk2019. We adjust call significance at Bonferroni adjusted threshold of *p_SMR_ <* 0.05 (a nominal *p_SMR_ <* 2.95 *∗* 10*^−^*^7^). This estimate assumes that the SNP used is not independently associated with DNAm and PD risk. As such, we report candidate CpG sites that show evidence of colocalization at a posterior probability *P* (*H*_4_) *>* 0.9, which helps to rule out instances whereby the candidate SNP in SMR only appears associated with both DNAm and PD risk due to LD.^23, 67, 70, 71^

## 3 Results

### 3.1 Best-fit Parkinson’s GRS show on-par prediction of disease status relative to the GRS showing the best performance in EWAS

Prior to EWAS, we assessed GRS performance in both our discovery sample (TERRE) and our replication sample (DIG-PD). In TERRE, we observed a maximum *R*^2^ = 0.0541 at *p <* 3.0 *∗* 10*^−^*^4^ in cross-sex samples, a maximum *R*^2^ = 0.102 at *p <* 0.034 in male samples, and a maximum *R*^2^ = 0.0696 at *p <* 3.5 *∗* 10*^−^*^4^ in female samples. This corresponds to total variation in PD explained of 2.56% in cross-sex samples, 4.72% in male samples, and 3.58% in female samples on the liability scale, which on par with the estimated 3.52 to 7.92% of variation explained by the loci in the most recent GWAS of PD status.^25^ At our EWAS threshold (i.e., *GRS_pT_* _=5*e*_*_−_*_8_, see Methods), we observed an *R*^2^ = 0.0121 in cross-sex samples, an *R*^2^ = 0.0273 in male samples, and an *R*^2^ = 0.0010 in female samples (also see Figure S6) which explain less variation in PD on the liability scale at 0.659% in cross-sex samples, 1.52% in male samples, and 0.0546% in female samples. Across all samples in DIG-PD, we observed a maximum *R*^2^ = 0.0361 at *p <* 5.0 *∗* 10*^−^*^8^ (our EWAS threshold), which corresponds to 2.89% variation in PD explained. Collectively, these results suggest that *GRS_pT_* _=5*e*_*_−_*_8_ is not universally predictive of PD, despite the fact that we ultimately detect an effect of this GRS on genome-wide DNAm.

Subsequently, we compared the predictive ability of each best-fit GRS to other PD risk factors (Table 1). TERRE is a unique PD sample that consists of French agricultural workers. As such, 46% of subjects (31 cases, 70 controls) reported regular use of at least one class of commercial-grade (Occupational) pesticide, and an additional 18% of subjects (15 cases, 25 controls) reported regular use of publicly available (Domestic) pesticides. However, we observed a positive trend in predicted PD risk with increasing GRS quartiles accounting for all other covariates (fourth quartile OR=2.60; 95% CI=[1.03, 6.55]; nominal *p <* 0.042; Table 1). The ratio of male to female PD patients was similar between TERRE and DIG-PD, and the subjects in the two samples had largely overlapping age ranges. However, DIG-PD had a higher proportion of cases and fewer subjects who were exposed to pesticides or had previously smoked tobacco. Nonetheless, in DIG-PD we observed a similar positive trend in predicted PD risk with increasing GRS quartiles (fourth quartile OR=2.84; 95% CI=[1.33, 6.09]; nominal *p <* 0.0072). Overall, the predictive performance of the best-fit GRS in both TERRE and DIG-PD is on par with what is seen in other studies, and the extent to which other covariates predict PD when accounting for the best-fit GRS is as expected.^72^

### 3.2 Parkinson’s GRS is associated strongly with DNAm in a 1.5 Mb window on chromosome 17

The first step of our EWAS was to determine the *GRS_pT_* that explained the greatest variability in DNAm across the genome. We iterated over 6000 pT (i.e., 6000 EWAS, Methods) and ultimately found that *pT* = 5 *∗* 10*^−^*^8^ yielded the most associations with DNAm overall, and for males and females separately (Figure S7A). This result aligns with that from an analysis of major depressive disorder in which the GRS that best predicted changes in DNAm was constructed at a lower pT than the GRS that best predicted disease status.^55^ Thus, we carried out all subsequent EWAS with a GRS made from SNPs associated with PD at a threshold of *p <* 5 *∗* 10*^−^*^8^ (*GRS_pT_* _=5*e*_*_−_*_8_).

Our EWAS identified 85 CpG sites (FDR *<* 0.05, difference of M-values > 1.5), of which 73 (86%) were within a 1.5 Mb window on chromosome 17 (Figure 2A, Supplementary Table SI). The remaining 10 associations distributed across chromosomes 2, 7, 10, 11, 16, and 18. In the 1.5 Mb window 22 CpG sites (22/85) annotated to *MAPT*, and 14 (14/85) annotated to *KANSL1*, two adjacent genes that play a role PD pathology and past GWAS have considered SNPs overlapping each gene to be a part of the same locus.^73–75^ In Figure 2B, we visualize known genes overlapping *MAPT* and *KANSL1*, and the correlation between DNAm at these CpG sites. The level of DNAm at GRS-associated CpG sites near the 5’ end of *MAPT*, are strongly correlated with one another, and negatively correlated with cg18228076, the top-associated site within *MAPT*. Considering this along with the observation that 6/9 CpG sites in the *MAPT* promoter have DNAm that decreases with an increase in *GRS_pT_* _=5*e*_*_−_*_8_, it is likely that higher genetic risk for PD, as measured by *GRS_pT_* _=5*e*_*_−_*_8_, results in decreased methylation of the promoter, and increased methylation in the gene body of *MAPT*. Additionally, we see that the top associated CpG site within *MAPT* (cg18228076) is within an enhancer region, and that DNAm is negatively correlated with the DNAm top CpG site in *KANSL1* (cg09860564) located in an intergenic region not overlapping *MAPT*, suggesting that *GRS_pT_* _=5*e*_*_−_*_8_ may have mirrored effect on DNAm patterns for sites located in these two genes.

**Figure 1:**
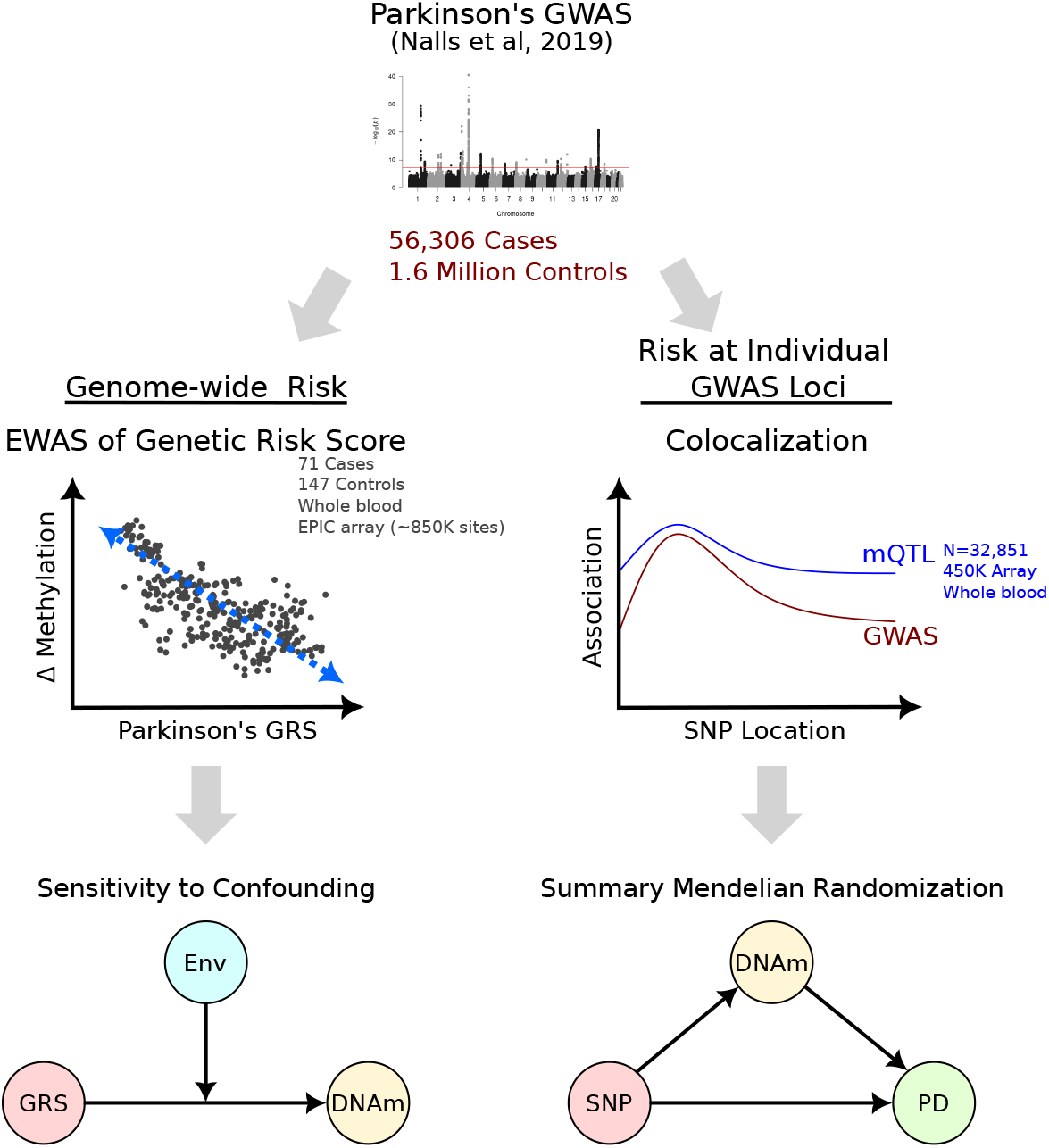
Overview of analysis. We take two approaches to explaining the effect of genetic risk for Parkinson’s disease (PD) on DNAm in blood. We first explained multi-locus genetic risk for PD in a genetic risk score (GRS), and then run an epigenome wide association study (EWAS) to discover its effect on DNAm, followed by a replication and sensitivity analysis. Second, we explained the effect of genetic risk for PD at individual loci using a colocalization analysis with blood mQTL, followed by a summary Mendelian randomization analysis to distinguish if any colocalized mQTL are consistent with DNAm mediating genetic risk for PD.

**Figure 2:**
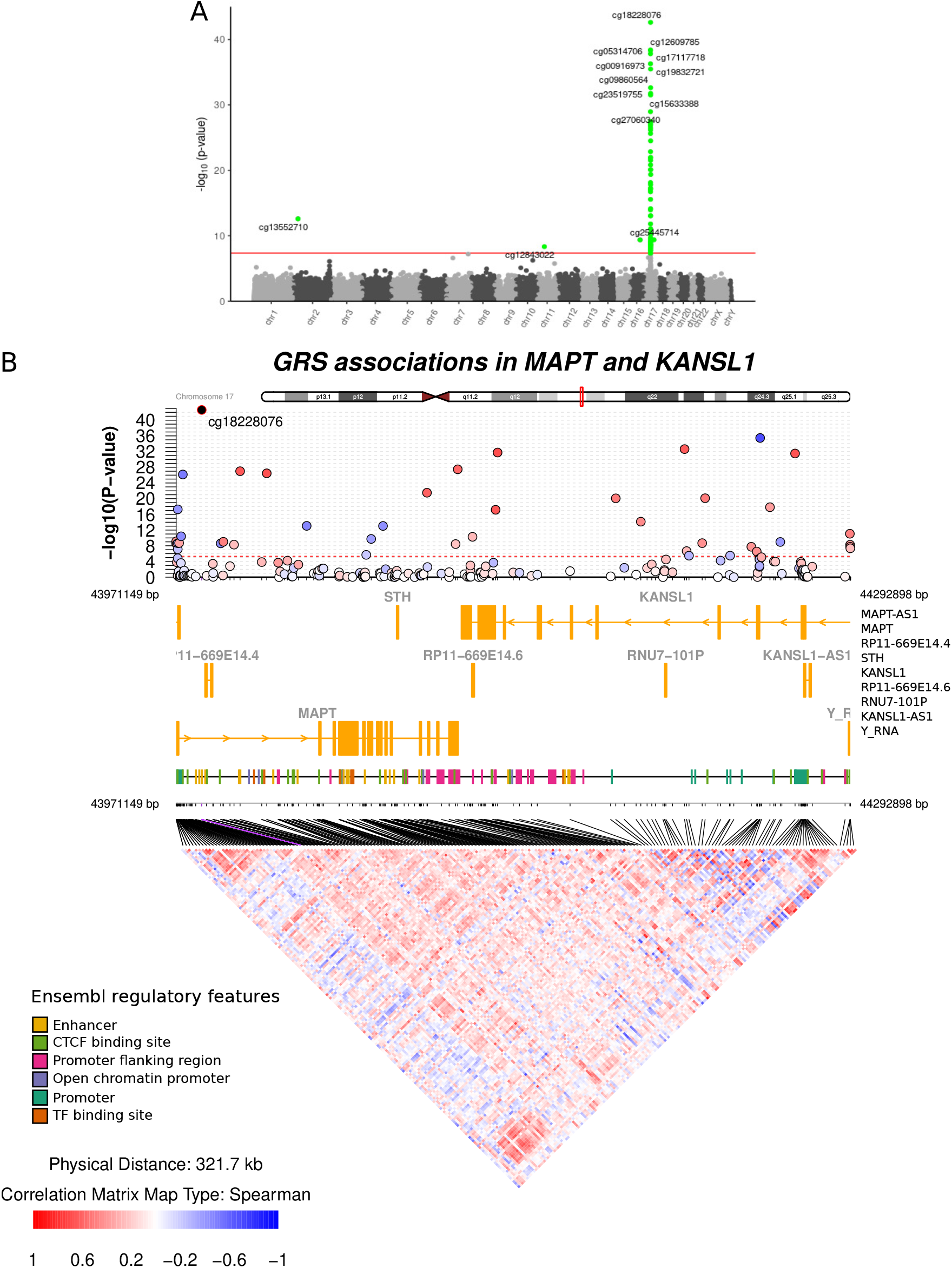
Parkinson’s GRS EWAS. **(A)**A Manhattan plot of the association between *GRS_pT_* _=5*e*_*_−_*_8_ and DNAm. We label the top 10 associations per chromosome with their identifier from the EPIC array. **(B)** A coMET plot displaying GRS-associated CpG sites annotated to either *MAPT* or *KANSL1*. Associated CpG sites are colored by their Spearman correlation with the top associated CpG site (cg18228076, within *MAPT*), and are aligned with known genes documented in the UCSC genome browser. All overlapping gene symbols are listed to the right of the plot. Known regulatory features from Ensembl are noted below this track. Lines are drawn between the position of each CpG site on the top of the graph to their position in bottom correlation matrix. The line corresponding to the top association (cg18228076) is highlighted in pink.

In a sensitivity analysis, we ran the EWAS using a GRS that excluded the six SNPs found on chromosome 17. The original and re-computed GRS we strongly correlated (Spearman’s *ρ* = 0.601 for in the cross-sex sample, *ρ* = 0.552 in males, *ρ* = 0.649 in females, Figure S8A,B). Most of all, we observed no difference in associated CpG sites, and we observed no difference in the number of associations with GRS at other p value thresholds.

To assess the robustness of our GRS-DNAm associations, we assessed replication in an independent sample (DIG-PD). We computed *GRS_pT_* _=5*e*_*_−_*_8_ for cross-sex, male, and female samples in DIG-PD and then analyzed the average log-fold change of DNAm M (Δ*M*) in cross-sex, male, and female samples separately (Figures 3A–C). Of 85 CpG sites associated with the *GRS_pT_* _=5*e*_*_−_*_8_ in TERRE cross-sex samples, 19 (22.4%) were also associated in DIG-PD cross-sex samples (*FDR <* 0.05), and effect of *GRS_pT_* _=5*e*_*_−_*_8_ at the 85 CpG sites showing association in TERRE were strongly correlated with effects in DIG-PD (Spearman’s *ρ* = 0.926, Figure 3A). In male and female samples there was less overlap across studies: nine CpG sites (15.3%) replicated with in males, and zero CpG sites replicated in females. However, the correlation between Δ*M* in TERRE and DIG-PD was fairly high in both sexes (*ρ* = 0.943 males and *ρ* = 0.596 females).

**Figure 3:**
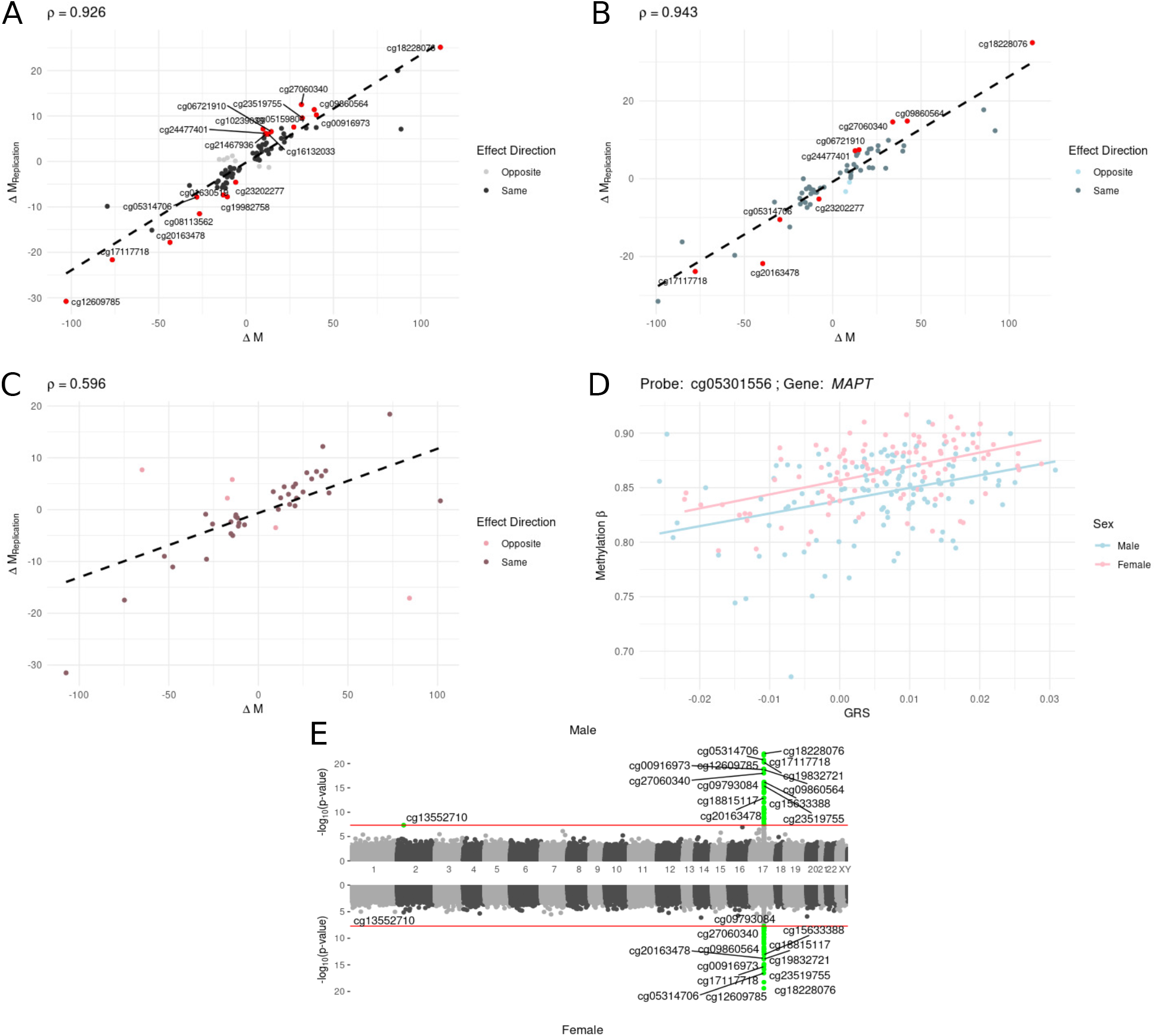
Replication and sensitivity to sex for GRS EWAS associations. Correlation between the average log-fold change in DNAm M-values in TERRE samples (Δ*M*) vs. DIG-PD (replication) samples (Δ*M_replication_*) **(A)** in cross-sex samples, **(B)** in male samples, and **(C)** in female samples. In red are the CpG sites with methylation significantly associated with the GRS in both TERRE and DIG-PD. **(D)** Methylation *β* values vs. cross-sex GRS at a CpG site significantly associated with sex (cg05301556, annotated to *MAPT*.). There is no evidence of interaction with sex (i.e., the slope in males is parallel to the slope in females). **(E)** A Miami plot of the association of *GRS_pT_* _=5*e*_*_−_*_8_ with DNAm in TERRE males vs. TERRE females. Associations significant at an *FDR <* 0.05 are highlighted green, with the top 10 associations per chromosome in either males or females labeled.

### 3.3 Parkinson’s GRS associates with DNAm independent of non-genetic factors

Next we observed that non-genetic variables relevant to PD, such as smoking status, head trauma, alcohol consumption, age, or exposure to pesticides, had little to no observable effect on DNAm when accounting for *GRS_pT_* _=5*e*_*_−_*_8_, nor did their interaction with *GRS_pT_* _=5*e*_*_−_*_8_ (Methods, Table S1). Thus, the non-genetic variables do not appear to moderate the relationship between *GRS_pT_* _=5*e*_*_−_*_8_ and DNAm.

Sex associated with 22 of 85 (25.9%, FDR *<* 0.05) CpG sites associated with the cross-sex GRS (Table S2). However, none of these CpG sites showed evidence of interaction with *GRS_pT_* _=5*e*_*_−_*_8_, indicating that the effect of *GRS_pT_* _=5*e*_*_−_*_8_ was similar in males and females, but with a different average DNAm level in each sex (see Figure 3D). We identified many of the same CpG sites associated with *GRS_pT_* _=5*e*_*_−_*_8_ in the cross-sex sample in sex-stratified analysis. In total, 59 CpG sites associated with *GRS_pT_* _=5*e*_*_−_*_8_ in males, 52 of which were associated with *GRS_pT_* _=5*e*_*_−_*_8_ in the cross-sex sample. Similarly, 41 CpG sites associated with *GRS_pT_* _=5*e*_*_−_*_8_ in females, 38 of which were associated with *GRS_pT_* _=5*e*_*_−_*_8_ in the cross-sex sample. Given this overlap, it was unsurprising that all 35 CpG sites associated with *GRS_pT_* _=5*e*_*_−_*_8_ in both males and females also associated with *GRS_pT_* _=5*e*_*_−_*_8_ in the cross-sex sample, which indicates that sex has little effect on GRS associations at these sites (Figures 3E, S7).

However, there were 24 CpG sites associated with *GRS_pT_* _=5*e*_*_−_*_8_ in either both the cross-sex sample and male samples, five of which exclusively associated with DNAm in male samples, and 16 of which were within the previously identified 1.5 Mb region on chromosome 17. Similarly, there were six CpG sites associated with either *GRS_pT_* _=5*e*_*_−_*_8_ in the cross-sex sample and female samples, three of which exclusively associated with DNAm in female samples. Although sex could play a role in the association between GRS and DNAm, we identified relatively few sites not also associated in a cross-sex sample of TERRE, and thus we were unable to detect convincing evidence of sex-specific effects in this study.

Other than sex and age, none of the covariates assessed in TERRE were measured in DIG-PD controls. As in TERRE, we observed little effect of these variables on the association of *GRS_pT_* _=5*e*_*_−_*_8_ with DNA. In DIG-PD, no CpG sites had DNAm associated (FDR *<* 0.05) with sex while accounting for *GRS_pT_* _=5*e*_*_−_*_8_, and a single CpG site, cg12798194, also showed a significant association with age when accounting for *GRS_pT_* _=5*e*_*_−_*_8_, with no significant interaction effect. Thus, the association of *GRS_pT_* _=5*e*_*_−_*_8_ with DNAm appears similarly invariant to some aspects of non-genetic variation in an independent sample.

### 3.4 Parkinson’s risk loci colocalize with different CpG sites than those associated with Parkinson’s GRS

We next assessed whether the effect of *GRS_pT_* _=5*e*_*_−_*_8_ on DNAm recapitulated the local effects of top PD loci on DNAm, or otherwise captured an independent, average genetic effect on DNAm. In total, 79 CpG sites had mQTL that colocalized with PD-associated genetic loci. *GRS_pT_* _=5*e*_*_−_*_8_ did not associate with DNAm at any of these sites. Unlike CpG sites where DNAm associated with *GRS_pT_* _=5*e*_*_−_*_8_, only 12 (15%) of these CpG sites are within chromosome 17. In fact, the majority of CpG sites with colocalized mQTL were within chromosomes 16 (23, or 29%) and 11 (18, or 23%) (Figure 4A, Supplementary Table SII). Within chromosome 17, only a single CpG site within the gene body of *MAPT* had an mQTL colocalized with PD risk (cg19276540). However, of the 73 CpG sites on chromosome 17 that associated with *GRS_pT_* _=5*e*_*_−_*_8_ only 35 were available on the 450K array and therefore available for colocalization analysis.

**Figure 4:**
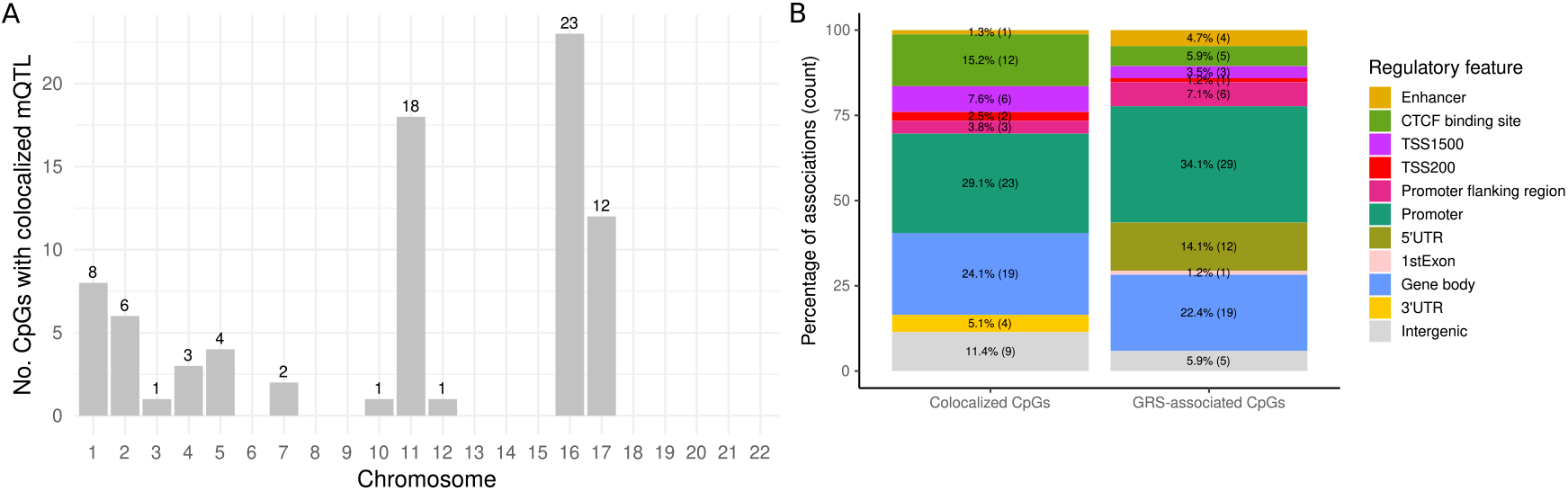
Colocalization between mQTL and individual PD GWAS loci. We computed colocalization of PD GWAS loci and mQTL from GoDMC at CpG sites within 1 Mb of each GWAS locus SNP. **(A)** The number of CpG sites with an mQTL colocalized with PD risk per chromosome. **(B)** The proportion of CpG sites from colocalization and GRS analysis annotated to regulatory features annotated by Ensembl, or otherwise annotated to gene regions as specified on Illumina’s EPIC array annotation: 3’ and 5’ UTR correspond to the 3’ and 5’ untranslated regions, TSS1500 and TSS200 correspond to a region within 1500 and 200 bp of the transcription start site, and gene body refers to a region within the gene excluding the first exon. The remaining CpG sites with no annotation are labeled as intergenic.

CpG sites with colocalized mQTL were similarly distributed in regulatory regions as compared to CpG sites associated with *GRS_pT_* _=5*e*_*_−_*_8_ (Figure 4B). The features we explored were generated from epigenomic information curated by Ensembl.^76^ CpG sites outside these regions were labeled using the EPIC array annotation. Briefly, the location of a CpG site relative to a regulatory feature can dictate the effect of its methylation on gene expression and other regulatory mechanisms, often as a result of methylation blocking the binding of regulatory proteins. For example, increased methylation at promoter sequences often represses transcription of the gene at that promoter and methylation of a CTCF binding site can inhibit longer range interactions between enhancer regions and promoters.^77, 78^ A smaller proportion of CpG sites with colocalized mQTL were promoter regions compared to CpG sites associated with *GRS_pT_* _=5*e*_*_−_*_8_ (23/79 or 29.1% of colocalized CpGs vs. 29/85 or 34.1% of GRS-associated CpGs). However, 41 GRS-associated CpG sites are only measured on the EPIC array, and thus not available in the colocalization analysis. Of 450K array sites, 23/41, or 56.1% of all 450K sites associated with *GRS_pT_* _=5*e*_*_−_*_8_, were within promoter regions. The proportion of colocalized CpG sites in gene bodies was similar to that of GRS-associated CpG sites (19/79 or 24.1% vs. 19/85 or 22.4% respectively). However, the proportion of GRS-associated CpG sites on the 450K array was 5/41, or 12.2%. Likewise, a larger proportion of colocalized CpG sites were within CTCF binding sites (12/79 or 15.2% vs. 5/85 or 5.9% of GRS-associated CpG sites, 2/41 or 4.9% of 450K sites). Thus, it is unclear whether the most common functions of CpG sites colocalized with PD risk differ from CpG sites associated with *GRS_pT_* _=5*e*_*_−_*_8_.

Genetic studies of PD suggest several other common loci that show a strong association with PD risk that did not show an association with blood DNAm in this study. For example, genetic studies detect association between loci within the *SNCA* gene, which encodes *α*-synuclein.^1, 79–81^ *SNCA* is of particular interest given its role in PD etiology, and DNAm in *SNCA* has been associated with DNAm in past studies conducted in blood and cerebral cortex.^82^ Our work considered three loci within the transcribed regions of *SNCA* during GRS analysis, and eight loci during mQTL-based analyses. Independently, each of the tag SNPs of these loci has a blood mQTL (Bonferroni-adjusted p *<* 0.05). Still, in colocalization analysis, we determined that the association between each of these loci with DNAm is likely independent of their association with PD status.

The previous colocalization analysis allowed us to rule out situations in which SNPs were associated with both DNAm and PD risk simply due to linkage (i.e., the SNP responsible for changes in DNAm was in linkage disequilibrium (LD) with the SNP that influences PD risk). Colocalization analysis, however, cannot distinguish between pleiotropy (i.e., the SNP influences DNA methylation and PD risk via two independent biological pathways) vs. causal association (i.e., the SNP has a causal effect on PD risk that is mediated by changes in DNAm). To distinguish between these two scenarios, we used summary Mendelian randomization (SMR).^69^ At CpG sites having mQTL overlapping PD GWAS loci, we first generated an estimate of the causal association of DNAm with PD using mQTL and GWAS summary statistics (Section 2.7). After generating estimates at all top mQTL, we called significant SMR associations at a Bonferroni-corrected *p <* 0.05. Of 175 CpG sites with a significant effect on PD risk (i.e., where top mQTL corresponded to a significant GWAS association), 25 a met colocalization criteria (*P* (*H*_4_ *>* 0.9) and with respect to a CpG site within 1Mb of a GWAS locus, Figure 5A, Supplementary Table SIII).

**Figure 5:**
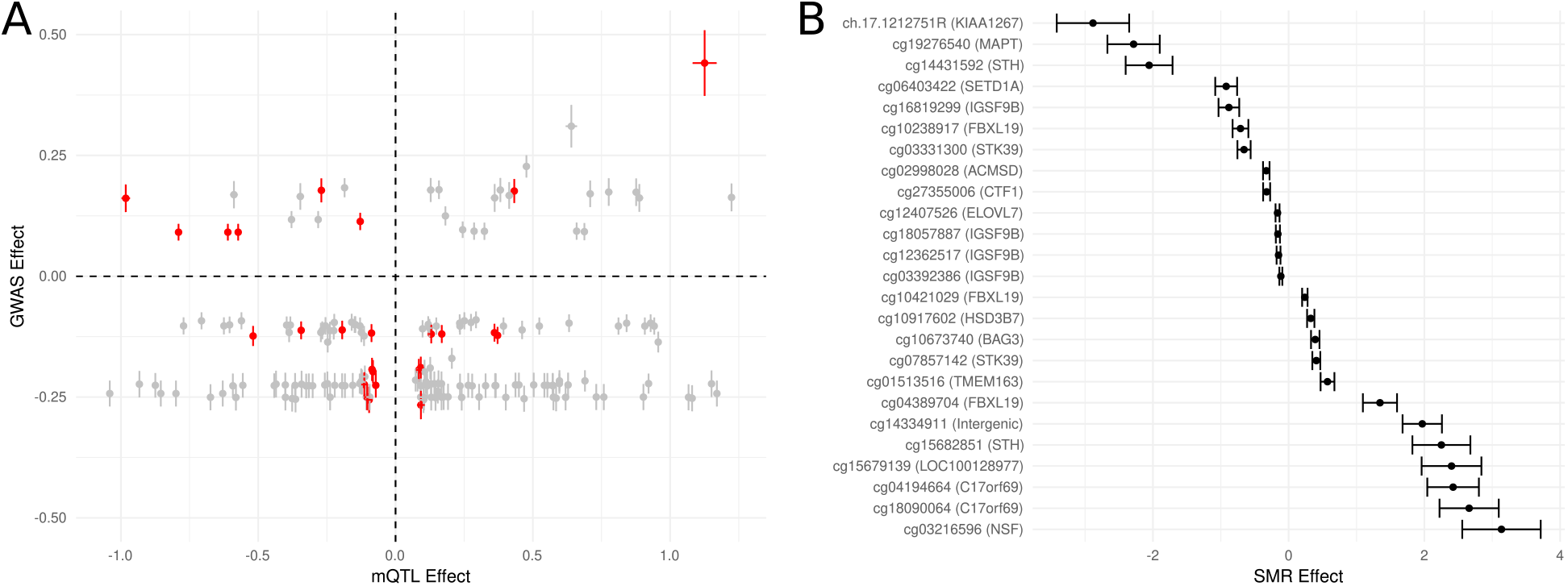
Summary Mendelian randomization for the effect of DNAm on PD risk. We computed summary Mendelian randomization for each mQTL in GoDMC matching a genome-wide significant SNP from the same independent GWAS used to compute GRS. **(A)** The mQTL vs. GWAS effect size of SMR estimates with a Bonferroni corrected *p <* 0.05. We mark estimates where the CpG site in question corresponded to an mQTL with *P* (*H*_4_) *>* 0.9 of colocalization in red, and show standard errors for each effect. For sites passing this cutoff, we estimate the effect of DNAm on PD risk as the ratio of GWAS and mQTL effect sizes, thus even if a SNP has large effects in both mQTL analysis and GWAS, the effect of DNAm on PD risk at a particular mQTL may be small. **(B)** We show the SMR effect for each significant SMR estimate at a CpG site with an mQTL colocalized with PD risk. Values greater than 0 indicate that an increase in DNAm at this site increases risk for PD, and likewise values less than 0 indicate that an increase in DNAm at this site decreases risk for PD. Additionally, we show the standard error of each of these estimates.

The nine CpG sites with the largest effect on PD risk were within chromosome 17 (Figure 5B), including one CpG site within the gene body of *MAPT* (cg19276540, an effect of *−*2.29 *±* 0.39, indicating that an increase in methylation at this site is associated with a decrease in PD risk). Additionally, a CpG site within the gene body of *BAG3* (cg10673740) stood out as having the largest GWAS and mQTL effect, although an increase methylation at this site leads to a smaller increase in PD risk (0.39 *±* 0.062). This gene can regulate the clearing of *α*-synuclein and MAPT protein.^83–85^ However, DNAm at these 25 CpG sites was not associated with *GRS_pT_* _=5*e*_*_−_*_8_. Thus, despite DNAm mediating the effect of genetic risk on PD at several sites on chromosome 17, this effect appears at distinct CpG sites from those with DNAm associated with *GRS_pT_* _=5*e*_*_−_*_8_.

## 4 Discussion

To date there are few CpG sites where DNAm appears associated with PD status (such as within *SLC7A11* and *CYP2E1*), and replication of these associations is low.^12, 86^ One possible reason for this is that case-control EWAS of PD are sensitive to the many genetic and non-genetic factors that influence both risk for PD and changes in DNAm. In order to address these concerns, we instead asked how genetic risk for PD influences DNAm in people with and without PD. Our approach builds on past studies showing that the association of genetic risk for a condition with DNAm is potentially less sensitive to confounders than the association between the condition and DNAm.^55, 59, 60^ Here, we assessed the influence of multi-locus PD GRS on DNAm in blood, evaluated whether associations were sensitive to major non-genetic risk factors for PD, and lastly compared GRS-associated sites to those affected by individual PD GWAS loci in SMR analysis. In the GRS experiment, we found multiple associations with DNAm in a single 1.5 Mb region on chromosome 17 that contains *MAPT*, and found that these associations were largely invariant to non-genetic variables available in the TERRE study, including known exposure to several pesticide families (Table S1). Using colocalization and SMR analyses, we similarly found a strong effect of genetic risk for PD on DNAm, but at distinct CpG sites. Taken together, these analyses suggest several regions where genetic risk for PD associates with DNAm in blood, and provide an annotation of non-coding genetic variants that influence PD risk.

We found 85 CpG sites where DNAm associated with *GRS_pT_* _=5*e*_*_−_*_8_. Many of these associations were on chromosome 17 (73/85 or 86%). The majority of these associations on chromosome 17 were with CpG sites annotated to *MAPT* (22 CpG sites) and a neighboring gene *KANSL1* (14 CpG sites). In a sensitivity analysis, we removed chromosome 17 variants from the GRS and re-tested association with DNAm. This had no effect on which CpG sites showed association, which suggests it is the sum of genetic effects that influence DNAm in this window, rather than the effect of individual GWAS loci within chromosome 17. Familial mutations in *MAPT* are associated with increased risk for frontotemporal lobar degeneration (FTLD) in patients with dementia and parkinsonism, as well as other neurodegenerative conditions like Alzheimer’s disease.^87–89^Given the associations we observe between *GRS_pT_* _=5*e*_*_−_*_8_, which excludes genetic loci on *MAPT*, and methylation within *MAPT*, it is possible that genetic risk conferred from loci external to *MAPT* can affect its function, and it is worth exploring if top PD risk loci are relevant to this process.

Past studies have grouped genetic variants in *KANSL1* into a single locus centered on *MAPT*.^25, 90^ However, recent work proposes that *KANSL1* could be involved in a separate pathway underlying PD etiology.^75^ Soutar *et al.* [75] demonstrated that knocking out *KANSL1* and *KAT8*, part of the non-specific lethal complex, in part decreases PINK1 activity in cell lines. In humans, decreased PINK1 activity associates with reduced synaptic activity in persons with PD. This mechanism would therefore involve interactions between genes on chromosomes 17 (*KANSL1*), 16 (*KAT8*), and 1 (*PINK1*). This coincides well with the pattern of association we observe with *GRS_pT_* _=5*e*_*_−_*_8_, in which top associated CpG sites in *MAPT* appear to have an opposite effect as those in *KANSL1*. Likewise, DNAm at top-associated CpG sites in *MAPT* do show weak correlation with DNAm at top-associated CpG sites across *KANSL1* (Figure 2B). Thus, the cluster of associations we observed in this 1.5 Mb window on chromosome 17 could coincide with several PD pathologies involving interactions of genes present across the genome.

We also felt it was important to explore whether non-genetic factors impacted our EWAS results given non-genetic variables associate with both PD and genome-wide DNAm. Genetic risk for PD, as estimated by twin studies, explains roughly 27% of PD risk, thus PD risk attributable to non-genetic factors and their interaction with genetic variation is potentially large (i.e., approaching 73%).^18, 24, 25^ For example, past work shows that occupational pesticide exposure can interact with genetic variants in *CYP2D* and *GSTT1* to affect PD risk, as observed in candidate gene case-control studies.^20, 91–93^ In this study, we were primarily concerned with the effect of PD GRS on genome-wide DNAm, and whether non-genetic risk factors for PD could confound our findings, especially given the difficulty of conducting case-control EWAS of PD.^12, 13^ Of the non-genetic variables analyzed in TERRE, including occupational pesticide exposure, only sex showed a significant association with DNAm after accounting for the *GRS_pT_* _=5*e*_*_−_*_8_. Despite sex associating with DNAm at 22 CpG sites, there was no evidence of interaction effects with *GRS_pT_* _=5*e*_*_−_*_8_. However, when stratifying our GRS analysis by sex, *GRS_pT_* _=5*e*_*_−_*_8_ uniquely associated with DNAm at five CpG sites in males and uniquely associated with DNAm at three CpG sites in females. This suggests sex could have a small influence on the relationship between genetic risk for PD and DNAm at these sites. Meanwhile, the finding that the majority (73 of 74) of non-genetic variables had no significant influence on the association of genetic risk for PD and DNAm aligns with both observations in GRS EWAS of other conditions and with previously noted lack of associations between PD GRS and non-genetic factors in the UK Biobank.^59, 60, 72^ Thus, while there is a potentially independent role of sex in the association between *GRS_pT_* _=5*e*_*_−_*_8_ and DNAm, it appears the influence of genetic PD risk factors for on DNAm outweighs the influence of non-genetic PD risk factors.

Lastly, colocalization and SMR analyses allowed us to further disentangle whether our EWAS associations were independent of individual genetic loci affecting DNAm. Of 79 blood mQTL colocalized with PD in this study, all were different from those identified in the GRS analysis, 12 (15%) were on chromosome 17, and only one associated with a CpG site annotated to *MAPT* (cg19276540). For comparison, Li *et al.* [94] conducted a transcriptome wide association study (TWAS) of PD, which summarizes how genetic effects on gene expression affect disease risk. Their top TWAS association in dorsolateral prefrontal cortex (brain) was in *MAPT*. This work also included colocalization analysis with QTL of gene expression, splicing, and DNAm measured in dorsolateral prefrontal cortex tissue, which ultimately detected colocalization with a splicing QTL within *MAPT*. Alternative analyses used imputed-methylation analysis to demonstrate that genetically regulated DNAm within *MAPT* shows strong association with Parkinson’s risk.^95^ Lastly, Nalls *et al.* [25] conducted SMR analysis using their GWAS meta-analysis, independent mQTL and eQTL available from various brain tissues and independent eQTL available in substantia nigra tissue from GTEx.^96–98^ Of the eQTL and mQTL they found that passed their mediation criteria, just four correspond to genes annotated to the 25 CpG sites with mQTL passing mediation criteria in our analysis: *MAPT* (brain eQTL) *SETD1A* (blood eQTL), *TMEM163* (brain and blood eQTL), and *IGSF9B* (blood mQTL, at a CpG site not prioritized here). Thus, it appears PD loci in and around *MAPT* affect multiple molecular traits attached to *MAPT* expression.

There are several limitations of this work to consider when interpreting results. First, the comparison between GRS-associated CpG sites and CpG sites with colocalized mQTL (from GoDMC) may be biased by differences in the EPIC vs. the 450K array. However, we found no GRS-associated sites when we limited our GRS EWAS to sites on the 450K array. Also, since GoDMC is one of the largest mQTL analyses in blood (N=32,851), with 248,607 independent *cis*-mQTL, we are confident that our list of colocalized mQTL is thorough with respect to sites on the 450K array.^23, 25^ Second, the *GRS_pT_* _=5*e*_*_−_*_8_, at 36 SNPs, only captures a small fraction of variation PD (0.659% on the liability scale), and the variation in PD explained via *GRS_pT_* _=3*e*_*_−_*_4_, the best-fit GRS in TERRE, is still shy of what is observed in larger cohorts (2.57% vs. between 6-8% on the liability scale).^25^ As such, the CpG sites associated with *GRS_pT_* _=5*e*_*_−_*_8_ are not associated with broad, genome-wide genetic risk for PD, but rather with top associated GWAS loci. Nonetheless, the association between *GRS_pT_* _=5*e*_*_−_*_8_ is highly replicated (at 19 sites), which is large considering that one of the largest EWAS of PD status had no replicated associations.^12^ Lastly, the EWAS here is conducted in primarily European subjects, using GRS constructed from primarily European GWAS summary statistics. Genetic association studies solely in subjects of European ancestry likely miss population-specific variation relevant to complex diseases, which greatly affects the accuracy of GRS in non-European populations.^99–101^ Thus, some of these associations may less relevant to PD in non-European samples.

In conclusion, PD GWAS have explained much of the underlying genetic risk of sporadic PD, and since many of these loci are non-coding, discovering the molecular consequences of PD loci is an important next step in understanding its etiology. In this study, we observed that a PD GRS primarily associated with DNAm on chromosome 17 in a 1.5 Mb window containing *MAPT*. Moreover, these associations outnumber what is observed in past EWAS of PD case-control status, and show strong replication in an independent cohort. Meanwhile, individual GWAS loci colocalized with 79 blood mQTL at different sets of CpG sites, distributed along different chromosomes. However, in SMR analysis, DNAm at nine of 25 CpG sites within chromosome 17 showed evidence of causal association with PD risk, including a site within the gene body of *MAPT*, emphasizing this location in both EWAS and in locus-specific analysis. Collectively, this analysis emphasizes that genetic risk for PD has a strong effect on DNAm in blood, with particularly strong effects found at CpG sites within a small section of chromosome 17. The strong associations in this study provide a starting point for future analyses on the role of epigenetic variation in PD etiology, and integrative genetic-epigenetic analysis will likely continue to be useful for gaining insight into the etiology of other complex traits.

## Conflict of Interest Statement

The authors declare that the research was conducted in the absence of any commercial or financial relationships that could be construed as a potential conflict of interest.

## Author Contributions

WC, JKD, and SM contributed to the conception and design of this study. WC performed statistical analysis. SLS performed all quality control for DNA methylation data. FA, CD, JS, SLS, AB, and the DIG-PD study group provided data curation. WC wrote the first draft of the manuscript. WC and SLS wrote sections of the manuscript. JC, SM, AE, MK, and JKD provided supervision. All authors contributed to manuscript revision, read, and approved the submitted version.

## Supporting information

Table SII

TableSIII

## Data Availability

The TERRE and DIGPD DNAm and genotyping data analyzed in the present study are subject to access restrictions via the European Union General Data Protection Regulation (GDPR) to maintain participant privacy. Requests for access can be directed to, including the proposed purpose for data use, and are subject to governance constraints and privacy restrictions. GWAS summary statistics are publicly available via https://drive.google.com/file/d/1FZ9UL99LAqyWnyNBxxlx6qOUlfAnublN/view?usp=sharing. GoDMC mQTL summary statistics are publicly available via http://fileserve.mrcieu.ac.uk/mqtl/assoc_meta_all.csv.gz. Analysis code is publicly available at https://github.com/wilcas/pd_grs_ewas_integration. The original contributions presented in the study are included in the article/supplementary material, further inquiries can be directed to the corresponding author.

## Acknowledgments

This project was supported by a British Columbia Children’s Hospital Research Institute Catalyst Grant. DNAm profiling in TERRE and DIG-PD was supported by the Canadian Institutes of Health Research (CIHR) [EGM-141897], the French National Research Agency [ANR-15-EPIG-0001], and The Federal Ministry of Education and Research [01KU1503A]. The DIG-PD study was supported by a grant from the French Ministry of Health (PHRC AOR0810), and a grant from the Agence nationale de sécurité du médicament et des produits de santé (ANSM 2013). The research leading to these results has received funding from the program “Investissement d’Avenir” ANR-10-IAIHU-06. Genotyping of TERRE cases and controls and of DIG-PD controls was performed as part of the Comprehensive Unbiased Risk Factor Assessment for Genetics and Environment in Parkinson’s Disease (COURAGE-PD) consortium supported by the JPND (https://www.neurodegenerationresearch.eu/initiatives/annual-calls-for-proposals/closed-calls/risk-factors-2012/risk-factor-call-results/courage-pd/). JKD is supported by a NARSAD Young Investigator Grant from the Brain and Behavior Research Foundation, and is a Michael Smith Health Research BC Scholar. SLS was supported by a CIHR Doctoral Award.

We would like to thank Chaini Konwar and Sarah M. Merrill for their advice in processing DNAm used in this study and Graham Boucher for suggestions and comments on this manuscript. We thank Badreddine Mohand Oumoussa and Abiba Doukani for their technical support at the P3S Post-Genomic Platform of Sorbonne University, and the DIG-PD Study Group for providing clinical data of DIG-PD.

## Data and code availability

The TERRE and DIGPD DNAm and genotyping data analyzed in the present study are subject to access restrictions via the European Union General Data Protection Regulation (GDPR) to maintain participant privacy. Requests for access can be directed to, including the proposed purpose for data use, and are subject to governance constraints and privacy restrictions. The research protocol of the TERRE study was approved by the ethics committee of Hôpital du Kremlin-Bicêtre, and all subjects provided written informed consent. The research protocol of the DIG-PD study was approved by the ethics committee of the University of Paris VI, and all subjects provided written informed consent. GWAS summary statistics are publicly available via https://drive.google.com/file/d/ 1FZ9UL99LAqyWnyNBxxlx6qOUlfAnublN/view?usp=sharing. GoDMC mQTL summary statistics are publicly available via http://fileserve.mrcieu.ac.uk/mqtl/assoc_meta_all.csv.gz. Analysis code is publicly available at https://github.com/wilcas/pd_grs_ewas_integration. The original contributions presented in the study are included in the article/supplementary material, further inquiries can be directed to the corresponding author.

## Notes

### Competing Interest Statement

The authors have declared no competing interest.

### Author Declarations

The research protocol of the TERRE study was approved by the ethics committee of Hôpital du Kremlin-Bicêtre, and all subjects provided written informed consent. The research protocol of the DIG-PD study was approved by the ethics committee of the University of Paris VI, and all subjects provided written informed consent.

