## Supplementary Material for "A Parkinson’s disease genetic risk score associates with blood DNAm on chromosome 17"

#### DIG-PD Study Group

**Steering committee:** Jean-Christophe Corvol, MD, PhD (Pitié-Salpêtrière Hospital, Paris, principal investigator of DIG-PD), Alexis Elbaz, MD, PhD (CESP, Villejuif, member of the steering committee), Marie Vidailhet, MD (Pitié-Salpêtrière Hospital, Paris, member of the steering committee), Alexis Brice, MD (Pitié-Salpêtrière Hospital, Paris, member of the steering committee and PI for genetic analysis) ;
**Statistical analyses:** Alexis Elbaz, MD, PhD (CESP, Villejuif, PI for statistical analyses), Fanny Artaud, PhD (CESP, Villejuif, statistician);
**Principal investigators for sites (alphabetical order):** Frédéric Bourdain, MD (CH Foch, Suresnes, PI for site), Jean-Philippe Brandel, MD (Fondation Rothschild, Paris, PI for site), Jean-Christophe Corvol, MD, PhD (Pitié-Salpêtrière Hospital, Paris, PI for site), Pascal Derkinderen, MD, PhD (CHU Nantes, PI for site), Franck Durif, MD (CHU Clermont-Ferrand, PI for site), Richard Levy, MD, PhD (CHU Saint-Antoine, Paris, PI for site), Fernando Pico, MD (CH Versailles, PI for site), Olivier Rascol, MD (CHU Toulouse, PI for site);
**Co-investigators (alphabetical order):** Anne-Marie Bonnet, MD (Pitié-Salpêtrière Hospital, Paris, site investigator), Cecilia Bonnet, MD, PhD (Pitié-Salpêtrière Hospital, Paris, site investigator), Christine Brefel-Courbon, MD (CHU Toulouse, site investigator), Florence Cormier-Dequaire, MD (Pitié-Salpêtrière Hospital, Paris, site investigator), Bertrand Degos, MD, PhD (Pitié-Salpêtrière Hospital, site investigator), Bérangère Debilly, MD (CHU Clermont-Ferrand, site investigator), Alexis Elbaz, MD, PhD (Pitié-Salpêtrière Hospital, Paris, site investigator), Monique Galitsky (CHU de Toulouse, site investigator), David Grabli, MD, PhD (Pitié-Salpêtrière Hospital, Paris, site investigator), Andreas Hartmann, MD, PhD (Pitié-Salpêtrière Hospital, Paris, site investigator), Stephan Klebe, MD (Pitié-Salpêtrière Hospital, Paris, site investigator), Julia Kraemmer, MD (Pitié-Salpêtrière Hospital, site investigator), Lucette Lacomblez, MD (Pitié-Salpêtrière Hospital, Paris, site investigator), Sara Leder, MD (Pitié-Salpêtrière Hospital, Paris, site investigator), Graziella Mangone, MD, PhD (Pitié-Salpêtrière Hospital, Paris, site investigator), Louise-Laure Mariani, MD (Pitié-Salpêtrière Hospital, Paris, site investigator), Ana-Raquel Marques, MD (CHU Clermont-Ferrand, site investigator), Valérie Mesnage, MD (CHU Saint Antoine, Paris, site investigator), Julia Muellner, MD (Pitié-Salpêtrière Hospital, Paris, site investigator), Fabienne Ory-Magne, MD (CHU Toulouse, site investigator), Violaine Planté-Bordeneuve, MD (Henri Mondor Hospital, Créteil, site investigator), Emmanuel Roze, MD, PhD (Pitié-Salpêtrière Hospital, Paris, site investigator), Melissa Tir, MD (CH Versailles, site investigator), Marie Vidailhet, MD (Pitié-Salpêtrière Hospital, Paris, site investigator), Hana You, MD (Pitié-Salpêtrière Hospital, Paris, site investigator);
**Neuropsychologists:** Eve Benchetrit, MS (Pitié-Salpêtrière Hospital, Paris, neuropsychologist), Julie Socha, MS (Pitié-Salpêtrière Hospital, Paris, neuropsychologist), Fanny Pineau, MS (Pitié-Salpêtrière Hospital, Paris, neuropsychologist), Tiphaine Vidal, MS (CHU Clermont-Ferrand, neuropsychologist), Elsa Pomies (CHU de Toulouse, neuropsychologist), Virginie Bayet (CHU de Toulouse, neuropsychologist);
**Genetic core:** Alexis Brice (Pitié-Salpêtrière Hospital, Paris, PI for genetic studies), Suzanne Lesage, PhD (INSERM, ICM, Paris, genetic analyses), Khadija Tahiri, PhD (INSERM, ICM, Paris, lab technician) Hélène Bertrand, MS (INSERM, ICM, Paris, lab technician), Graziella Mangone, MD, PhD (Pitié-Salpêtrière Hospital, Paris, genetic analyses);
**Sponsor activities and clinical research assistants:** Alain Mallet, PhD (Pitié-Salpêtrière Hospital, Paris, sponsor representative), Coralie Villeret (Hôpital Saint Louis, Paris, Project manager), Merry Mazmanian

(Pitié-Salpêtrière Hospital, Paris, project manager), Hakima Manseur (Pitié-Salpêtrière Hospital, Paris, clinical research assistant), Mostafa Hajji (Pitié-Salpêtrière Hospital, Paris, data manager), Benjamin Le Toullec, MS (Pitié-Salpêtrière Hospital, Paris, clinical research assistant), Vanessa Brochard, PhD (Pitié-Salpêtrière Hospital, Paris, project manager), Monica Roy, MS (CHU de Nantes, clinical research assistant), Isabelle Rieu, PhD (CHU Clermont-Ferrand, clinical research assistant), Stéphane Bernard (CHU Clermont-Ferrand, clinical research assistant), Antoine Faurie-Grepon (CHU de Toulouse, clinical research assistant).

### S1 Supplementary Methods

#### S1 Sex-stratified genetic risk scoring

Given the slight sex difference in genetic architecture of PD, we also compared sex-specific genetic risk to sex-agnostic genetic risk for PD in male vs. female samples in TERRE. We used publicly available sex-stratified GWAS summary statistics from[1], which contains a subset of samples in[2], to construct a male and female GRS. Again, we observed different degrees of fit to Parkinson's status across the same $p$-value thresholds (Figure S5C). We observed the same trends in optimal $R^2$ for the female GRS vs. the cross-sex GRS: the female GRS was close in fit to the cross-sex GRS ($R^2 = 0.0659$). The male GRS performed worse ($R^2 = 0.0221$) than the female GRS and was not significantly associated with PD status ($p < 0.33$). 59.8% of SNPs selected for the cross-sex GRS were present in the male GWAS GRS vs. 40.5% of SNPs present in the female GWAS GRS at a threshold of $p < 5 * 10^{-8}$. This analysis suggests some minor variability in cross-sex GRS performance with sex in TERRE. However, the extent to which using sex-stratified GRS could impact our results is still ambiguous given the poor fit of a male GWAS derived GRS to PD status in males, and thus we opted to not use sex-stratified GWAS results in our downstream analyses.

#### S2 Genome-wide cis-mQTL mapping

As a sensitivity analysis, we computed the association between DNAm beta values and imputed genotype dosage of each SNP $i$ within 75kb of each CpG site on the EPIC array using linear regression as implemented in `matrixEQTL`[3]. The majority of cis-mQTL appear within 50kb of the transcription start site in blood and several other human tissues[4]. We chose to extend this window by an additional 50kb total in order to include additional mQTL in our analysis. mQTL were computed using the following linear model in both TERRE and DIG-PD:

$$DNAm \sim intercept + SNP_i + sex + Age + genotypingPC_{\{1...3\}} + DNAmPC_{\{1...10\}} \quad (1)$$

Due to space restrictions, we only saved all associations with a nominal $p < 0.25$. We assessed replication of TERRE mQTL in GoDMC, using $\pi_1$ statistics on P values normalized by this threshold, which estimates the proportion of mQTL in TERRE with non-null effects in DIG-PD[5,6]. In TERRE, we detected 71,875 CpG sites with an mQTL significant at a Bonferroni adjusted $p < 0.05$, not accounting for correlation between CpG sites. These mQTL showed high replication in mQTL computed in GoDMC ($\pi_1 = 0.95$).

### S2 Supplementary Figures and Tables

#### S1 Figures

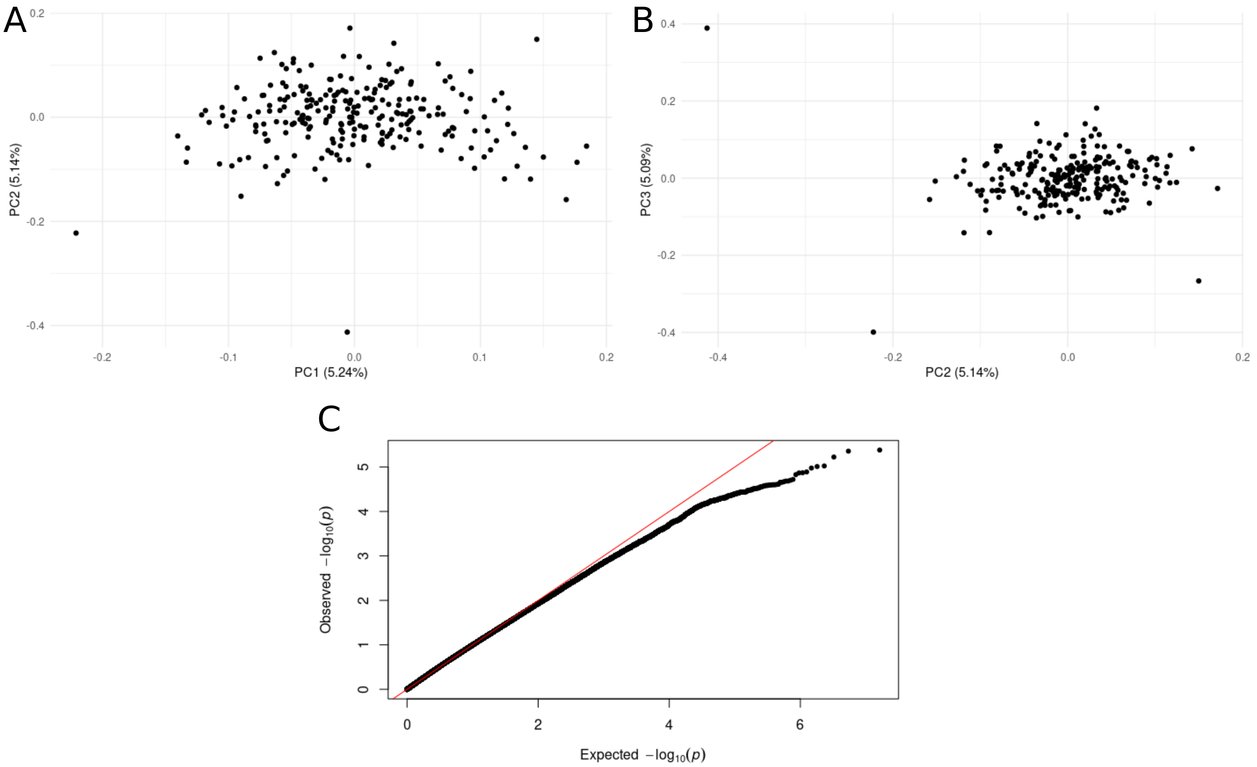


**Figure S1. Genotyping quality control in TERRE samples.** After applying quality control measures to TERRE, we computed genotyping principal components and plotted subjects along **(A)**components one and two, and **(B)** components two and three. **(C)** Expected vs. observed association between genotype and Parkinson's status in TERRE accounting for covariates.

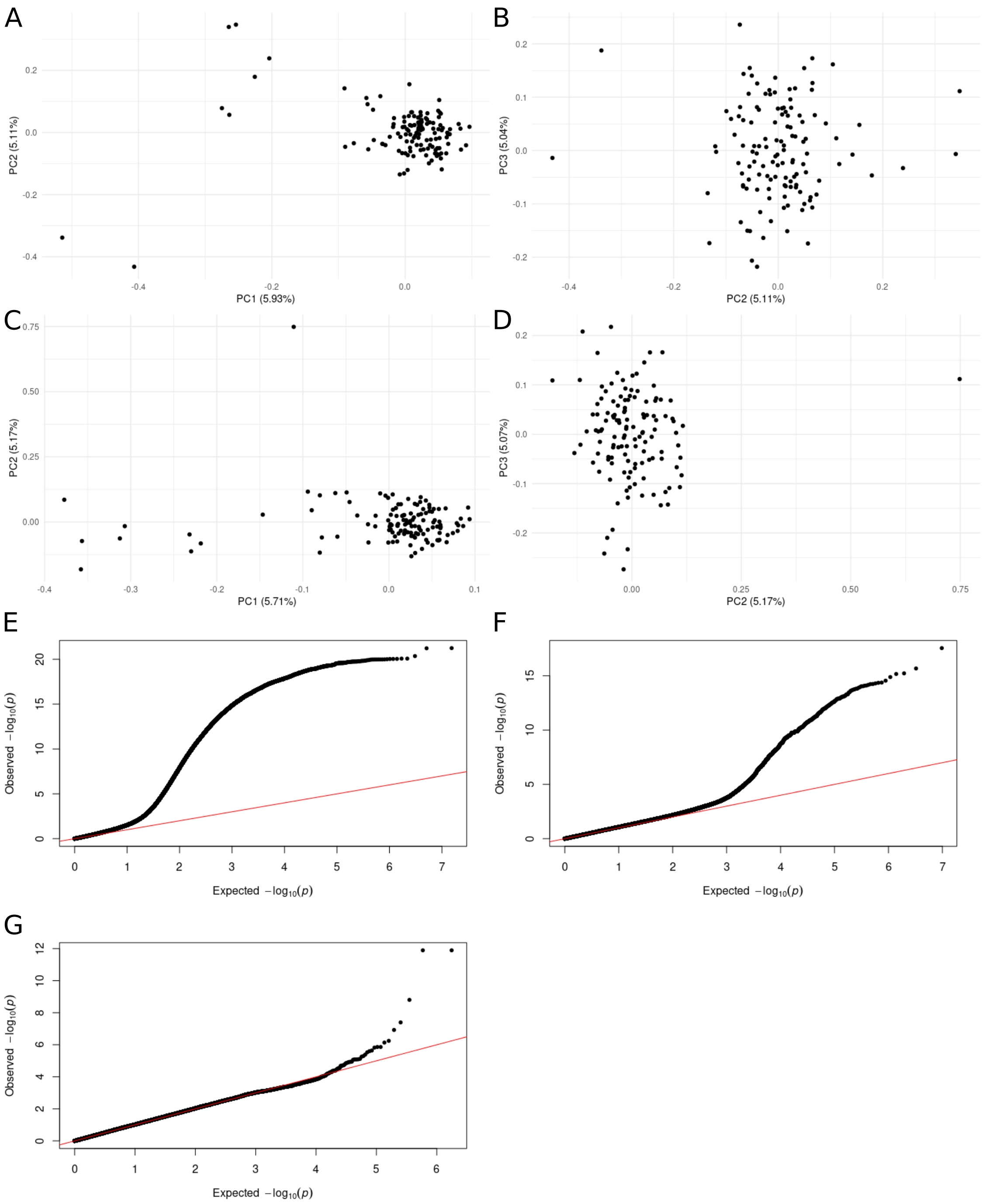


**Figure S2. Genotyping quality control in DIG-PD.** We applied quality control measures separately to cases and controls in DIG-PD, including the computation of genotyping PCs to account for cryptic relatedness, plotting samples along PCs follows: **(A)** first and second components for cases, **(B)** second and third components for cases, **(C)** first and second components for controls, and **(D)** second and third components for controls. We checked the association between imputed genotype dosage and Parkinson's status for inflation in samples merged after filtering at different imputation quality thresholds: **(E)** $R2 < 0.3$, **(F)** $R2 < 0.8$, and **(G)** $R2 < 0.9$.

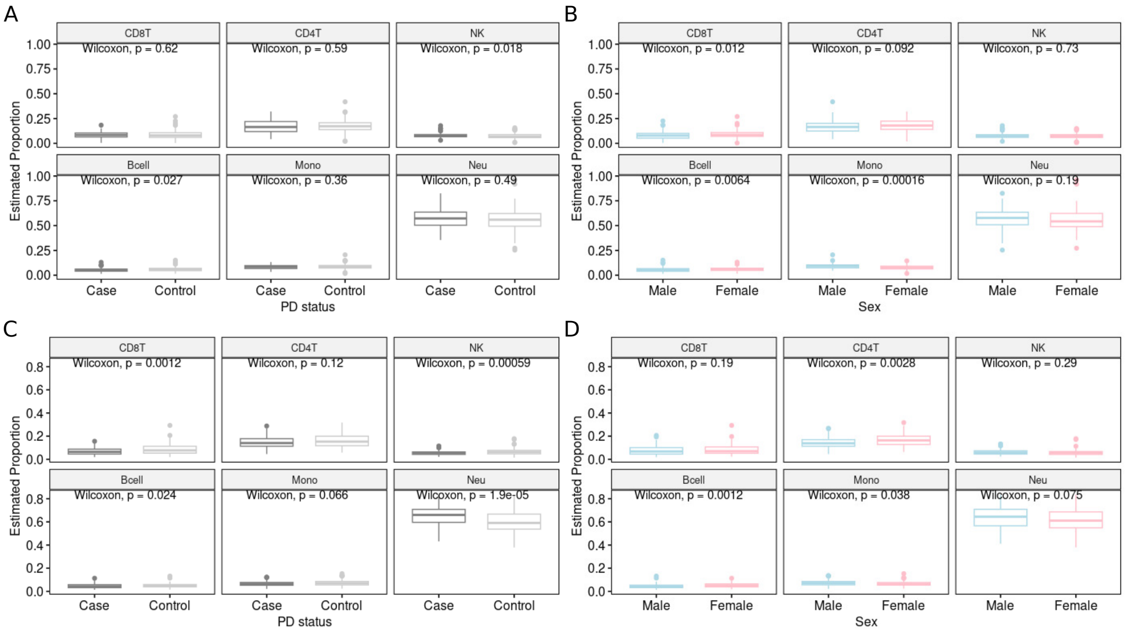

**Figure S3. Differences between cell type proportion across sex and PD case-control status in TERRE and DIG-PD** We tested differences in estimated immune cell proportions using a two-sided Wilcoxon test. **(A)** PD cases vs. controls in TERRE. **(B)** Male vs. female samples in TERRE. **(C)** PD cases vs. controls in DIG-PD. **(D)** Male vs. female samples in DIG-PD.

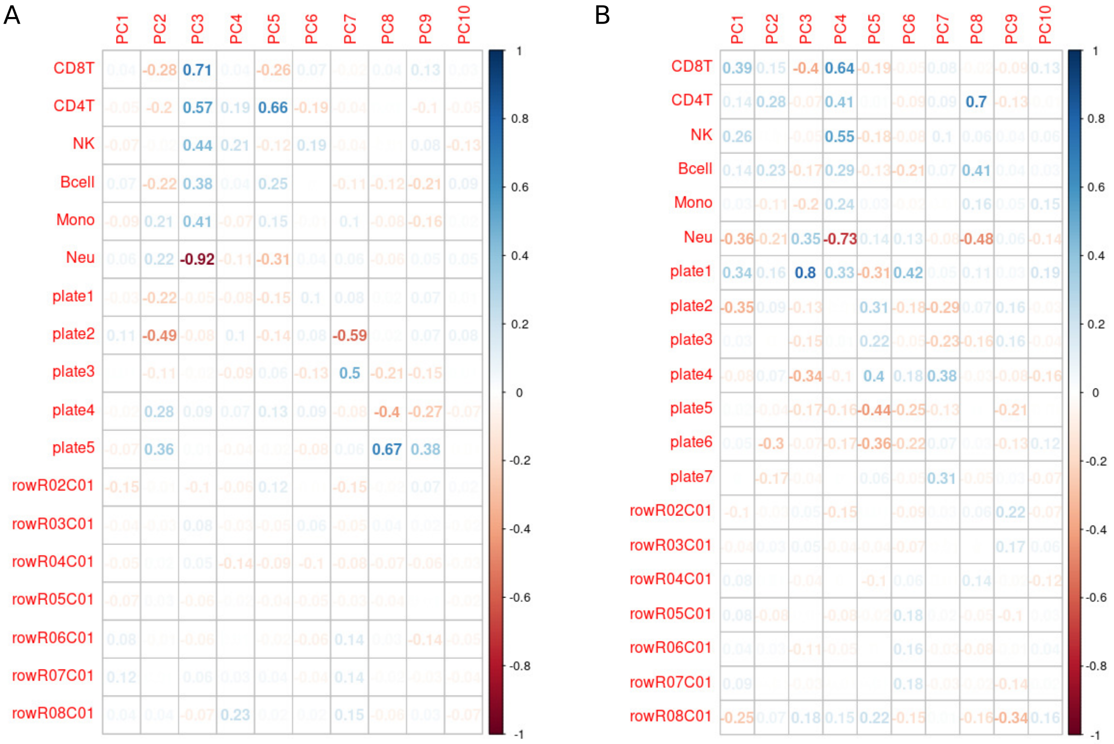

**Figure S4. Correlation between the top 10 principal components of DNAm and estimated immune cell type proportions and DNAm batch variables (plate and row). (A)** Correlations in TERRE samples. **(B)** Correlations in DIG-PD samples.

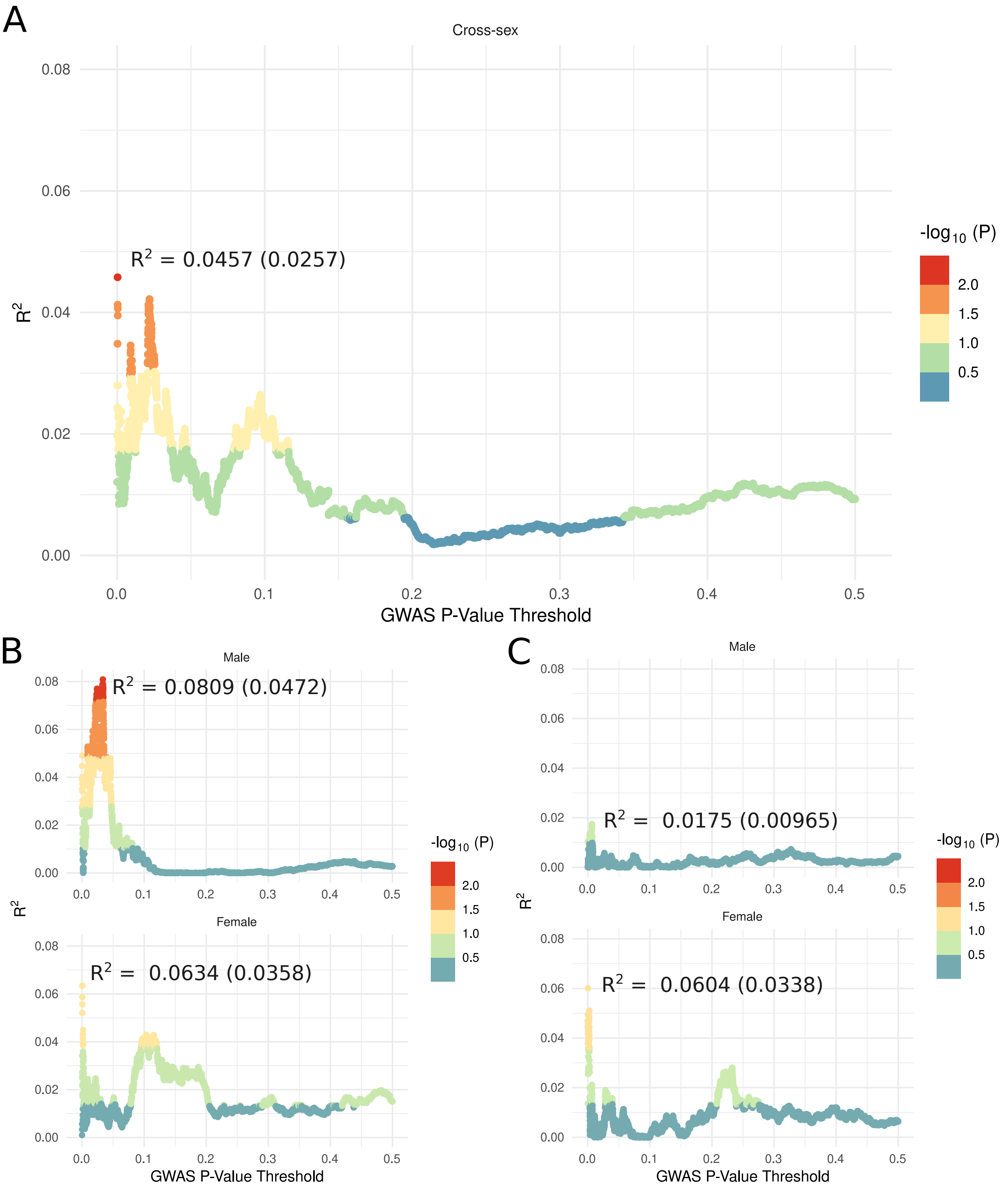


**Figure S5. Generating polygenic scores for cross-sex, male, and female samples in TERRE.** Adjusted $R^2$ values (on the sample scale) for GRS computed with cross-sex GWAS summary statistics vs. $p$-value thresholds for TERRE **(A)** cross-sex samples, and **(B)** male and female samples. **(C)** Adjusted $R^2$ values (on the sample scale) for GRS computed with male and female stratified GWAS summary statistics on male and female samples respectively. The largest adjusted $R^2$ values for each GRS are displayed on the sample scale, with $R^2$ on the liability scale in parentheses.

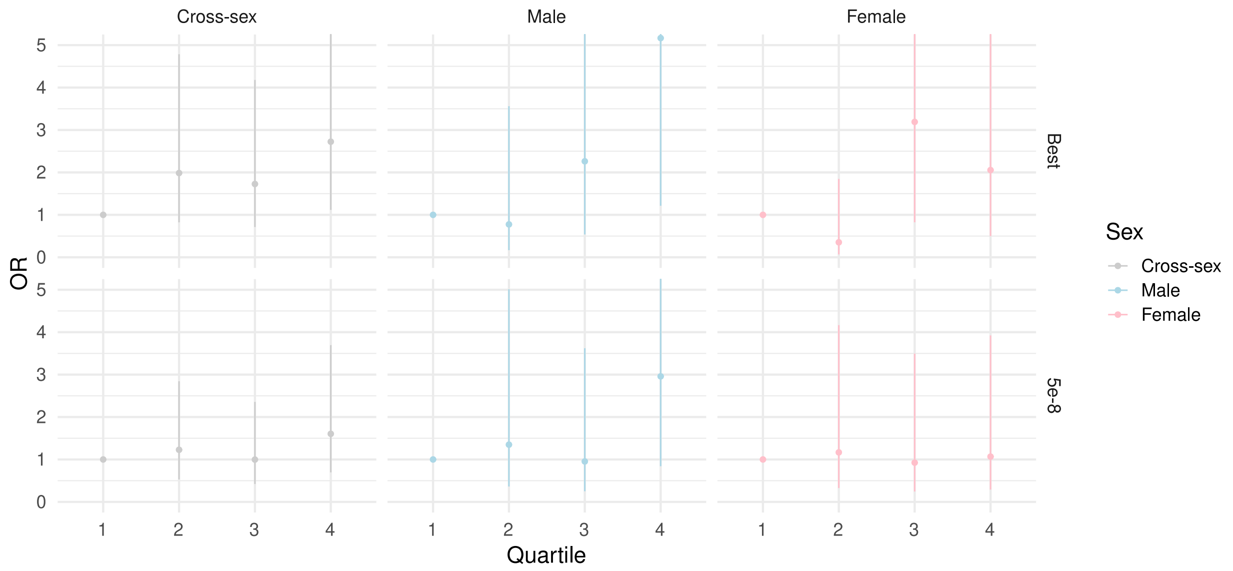


**Figure S6. Odds ratio of Parkinson's status relative to first genetic risk score quartile in TERRE.** In the top row, we display the odds ratio of PD status and its 95% confidence interval for the GRS with the highest adjusted $R^2$ value in the cross-sex sample and male and female samples respectively. In the bottom row, we show the odds ratio of PD status and 95% confidence interval for the $GRS_{pT=5e-8}$. GRS were constructed using cross-sex GWAS summary statistics. We report estimates from a multivariate model using the same covariates used in EWAS.

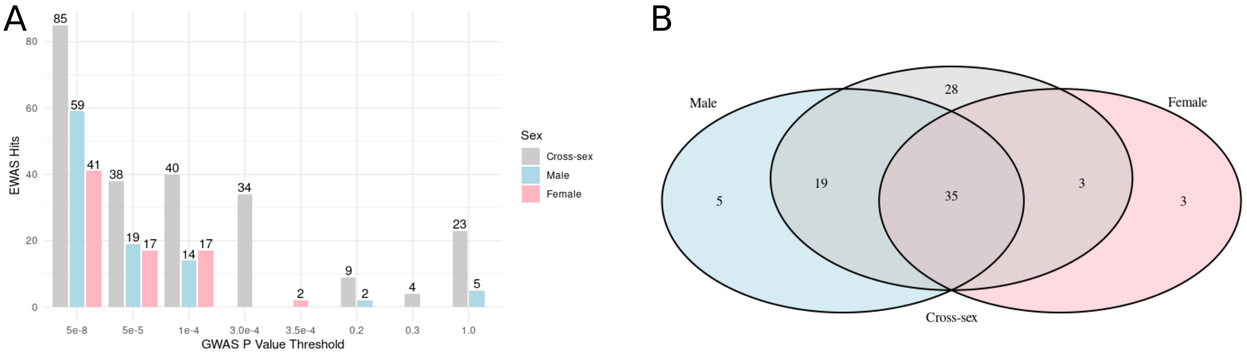


**Figure S7. CpG sites associated with Parkinson's genetic risk scores and overlap in sites. (A)** Association between $GRS$ with DNAm in TERRE at various $p$-value thresholds for cross-sex, male, and female samples. **(B)** Overlap in CpGs with DNAm associated with $GRS_{pT=5e-8}$ at an $FDR < 0.05$ in cross-sex, male and female samples.

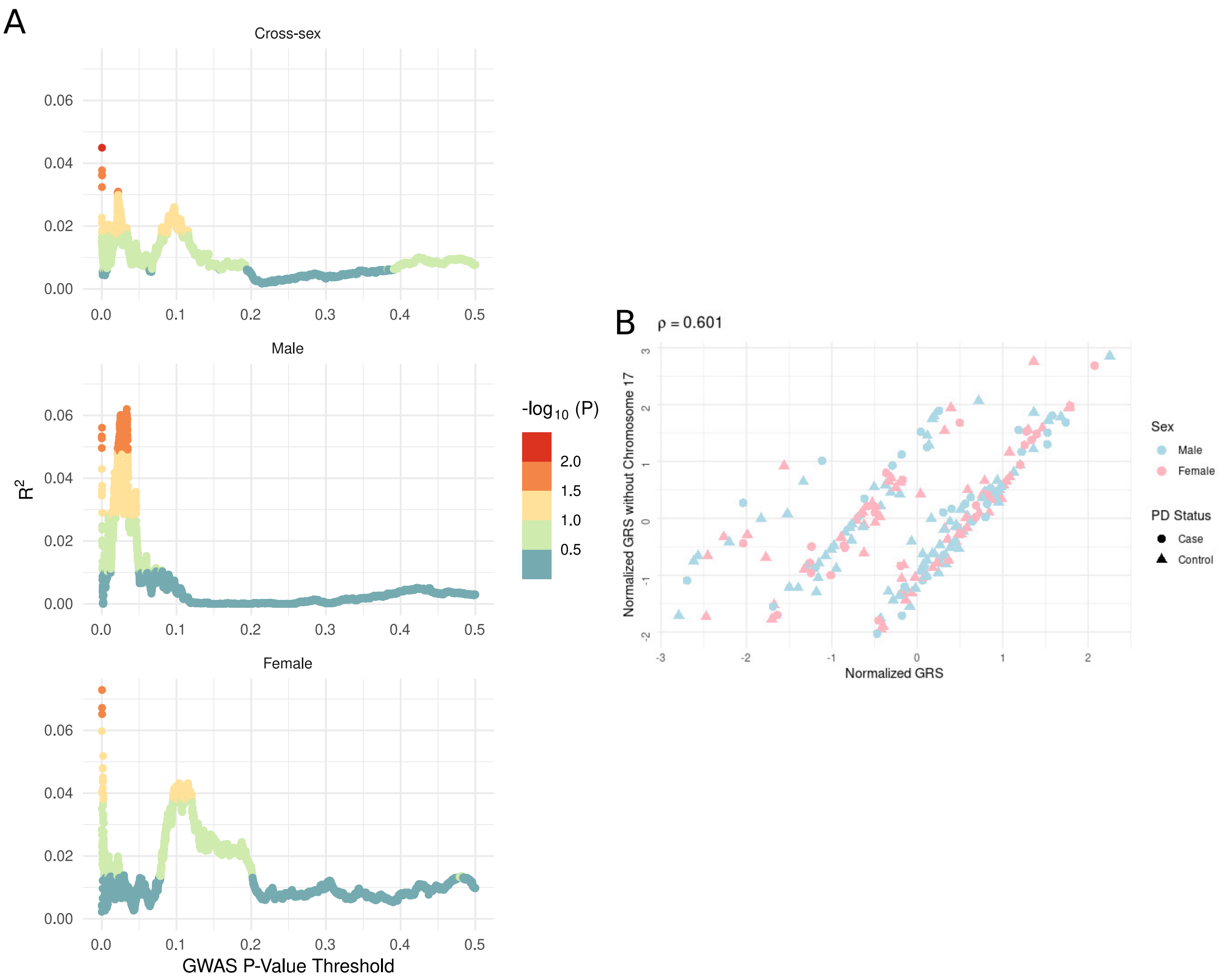


**Figure S8. Comparing GRS built with cross-sex GWAS summary statistics with and without SNPs on Chromosome 17(A)**Adjusted $R^2$ values for cross-sex, male, and female GRS computed on cross-sex GWAS summary statistics over SNPs associated with Parkinson's status at various $p$-value thresholds, excluding SNPs on chromosome 17.**(B)** Normalized GRS vs. normalized GRS built without chromosome 17 for cross-sex GWAS SNPs significant at $p < 5 * 10^{-8}$. We display Spearman's $\rho$ between the two scores. Bands appear as the scores differ by six SNPs in strong LD (36 SNPs vs. 30 SNPs when excluding SNPs on chromosome 17).

#### S2 Tables

**Table S1. Proportion of TERRE samples exposed to pesticides.** Estimates for each subject are average binary exposure across 10 imputations.

| | Case | Control | Total |
|---|---|---|---|
| | (N=72) | (N=139) | (N=211) |
| **Insecticides** | | | |
| Mean (SD) | 0.44 (± 0.50) | 0.41 (± 0.49) | 0.42 (± 0.49) |
| Missing | 0 (0%) | 1 (0.7%) | 1 (0.5%) |
| **Fungicides** | | | |
| Mean (SD) | 0.47 (± 0.50) | 0.38 (± 0.49) | 0.41 (± 0.49) |
| Missing | 0 (0%) | 1 (0.7%) | 1 (0.5%) |
| **Herbicides** | | | |
| Mean (SD) | 0.44 (± 0.50) | 0.38 (± 0.49) | 0.40 (± 0.49) |
| Missing | 1 (1.4%) | 0 (0%) | 1 (0.5%) |
| **Arsenic** | | | |
| Mean (SD) | 0.10 (± 0.29) | 0.064 (± 0.23) | 0.077 (± 0.25) |
| Missing | 0 (0%) | 1 (0.7%) | 1 (0.5%) |
| **Sulfur** | | | |
| Mean (SD) | 0.39 (± 0.48) | 0.33 (± 0.46) | 0.35 (± 0.47) |
| Missing | 0 (0%) | 3 (2.2%) | 3 (1.4%) |
| **Copper** | | | |
| Mean (SD) | 0.40 (± 0.48) | 0.35 (± 0.47) | 0.37 (± 0.47) |
| Missing | 0 (0%) | 2 (1.4%) | 2 (0.9%) |
| **Mercury** | | | |
| Mean (SD) | 0.038 (± 0.14) | 0.051 (± 0.17) | 0.046 (± 0.16) |
| **Sodium perchlorate** | | | |
| Mean (SD) | 0.041 (± 0.18) | 0.033 (± 0.17) | 0.036 (± 0.17) |
| Missing | 1 (1.4%) | 0 (0%) | 1 (0.5%) |
| **Oils** | | | |
| Mean (SD) | 0.014 (± 0.12) | 0.024 (± 0.15) | 0.020 (± 0.14) |
| Missing | 1 (1.4%) | 0 (0%) | 1 (0.5%) |
| **Organochlorine (Insecticides)** | | | |
| Mean (SD) | 0.31 (± 0.45) | 0.29 (± 0.44) | 0.30 (± 0.44) |
| Missing | 0 (0%) | 1 (0.7%) | 1 (0.5%) |
| **Organophosphorus (Insecticides)** | | | |
| Mean (SD) | 0.28 (± 0.41) | 0.23 (± 0.39) | 0.25 (± 0.40) |
| Missing | 0 (0%) | 1 (0.7%) | 1 (0.5%) |
| **Carbamate (Insecticides)** | | | |
| Mean (SD) | 0.11 (± 0.27) | 0.13 (± 0.32) | 0.13 (± 0.30) |
| Missing | 2 (2.8%) | 1 (0.7%) | 3 (1.4%) |

**Table S1 – continued from previous page**

| | Case | Control | Total |
|---|---|---|---|
| | (N=72) | (N=139) | (N=211) |
| **Pyrethroid (Insecticides)** | | | |
| Mean (SD) | 0.15 (± 0.34) | 0.18 (± 0.37) | 0.17 (± 0.36) |
| Missing | 2 (2.8%) | 1 (0.7%) | 3 (1.4%) |
| **Urea (Insecticides)** | | | |
| Mean (SD) | 0.014 (± 0.12) | 0.014 (± 0.12) | 0.014 (± 0.12) |
| **Sulfone (Insecticides)** | | | |
| Mean (SD) | 0.014 (± 0.12) | 0 (± 0) | 0.0047 (± 0.069) |
| **Natural (Insecticides)** | | | |
| Mean (SD) | 0 (± 0) | 0.022 (± 0.15) | 0.014 (± 0.12) |
| Missing | 2 (2.8%) | 0 (0%) | 2 (0.9%) |
| **Acaricide** | | | |
| Mean (SD) | 0 (± 0) | 0.020 (± 0.13) | 0.013 (± 0.11) |
| Missing | 2 (2.8%) | 0 (0%) | 2 (0.9%) |
| **Carbamate or dithiocarbamate (Fungicides)** | | | |
| Mean (SD) | 0.29 (± 0.44) | 0.22 (± 0.40) | 0.24 (± 0.41) |
| Missing | 1 (1.4%) | 1 (0.7%) | 2 (0.9%) |
| **Aromatic or dinitrophenol (Fungicides)** | | | |
| Mean (SD) | 0.017 (± 0.12) | 0.033 (± 0.17) | 0.027 (± 0.15) |
| Missing | 1 (1.4%) | 1 (0.7%) | 2 (0.9%) |
| **Amide (Fungicides)** | | | |
| Mean (SD) | 0.086 (± 0.27) | 0.061 (± 0.23) | 0.070 (± 0.24) |
| Missing | 2 (2.8%) | 2 (1.4%) | 4 (1.9%) |
| **Imide (Fungicides)** | | | |
| Mean (SD) | 0.072 (± 0.25) | 0.065 (± 0.24) | 0.067 (± 0.24) |
| Missing | 1 (1.4%) | 2 (1.4%) | 3 (1.4%) |
| **Diazine (Fungicides)** | | | |
| Mean (SD) | 0.044 (± 0.19) | 0.050 (± 0.19) | 0.048 (± 0.19) |
| Missing | 2 (2.8%) | 2 (1.4%) | 4 (1.9%) |
| **Triazine or triazole (Fungicides)** | | | |
| Mean (SD) | 0.10 (± 0.29) | 0.089 (± 0.27) | 0.093 (± 0.28) |
| Missing | 1 (1.4%) | 1 (0.7%) | 2 (0.9%) |
| **Morpholine (Fungicides)** | | | |
| Mean (SD) | 0.050 (± 0.21) | 0.025 (± 0.13) | 0.033 (± 0.16) |
| Missing | 0 (0%) | 2 (1.4%) | 2 (0.9%) |
| **Imidazole (Fungicides)** | | | |
| Mean (SD) | 0.015 (± 0.12) | 0.027 (± 0.15) | 0.023 (± 0.14) |
| **Piperidine (Fungicides)** | | | |

**Table S1 – continued from previous page**

| | Case | Control | Total |
|---|---|---|---|
| | (N=72) | (N=139) | (N=211) |
| Mean (SD) | 0.028 (± 0.17) | 0.0022 (± 0.025) | 0.011 (± 0.099) |
| **Phosphonate (Fungicides)** | | | |
| Mean (SD) | 0 (± 0) | 0.036 (± 0.19) | 0.024 (± 0.15) |
| **Anthraquinone (Bird repellent)** | | | |
| Mean (SD) | 0.040 (± 0.16) | 0.055 (± 0.18) | 0.050 (± 0.17) |
| **Oxine-copper (Fungicides)** | | | |
| Mean (SD) | 0.056 (± 0.19) | 0.067 (± 0.21) | 0.063 (± 0.20) |
| Missing | 0 (0%) | 1 (0.7%) | 1 (0.5%) |
| **Other (Fungicides)** | | | |
| Mean (SD) | 0 (± 0) | 0.015 (± 0.10) | 0.010 (± 0.085) |
| Missing | 1 (1.4%) | 1 (0.7%) | 2 (0.9%) |
| **Carbamate (Herbicides)** | | | |
| Mean (SD) | 0.029 (± 0.17) | 0.049 (± 0.20) | 0.042 (± 0.19) |
| Missing | 0 (0%) | 1 (0.7%) | 1 (0.5%) |
| **Dinitrophenol or Diphenyl ether or Benzonitrile or Aryloxyphenoxypropionic (Herbicides)** | | | |
| Mean (SD) | 0.28 (± 0.43) | 0.17 (± 0.35) | 0.21 (± 0.38) |
| Missing | 1 (1.4%) | 0 (0%) | 1 (0.5%) |
| **Phenoxyacetic (Herbicides)** | | | |
| Mean (SD) | 0.33 (± 0.47) | 0.27 (± 0.44) | 0.29 (± 0.45) |
| Missing | 1 (1.4%) | 1 (0.7%) | 2 (0.9%) |
| **Benzoic acid (Herbicides)** | | | |
| Mean (SD) | 0.033 (± 0.17) | 0.050 (± 0.19) | 0.045 (± 0.19) |
| **Picolinic acid (Herbicides)** | | | |
| Mean (SD) | 0.13 (± 0.31) | 0.12 (± 0.30) | 0.12 (± 0.30) |
| Missing | 0 (0%) | 1 (0.7%) | 1 (0.5%) |
| **Diazine (Herbicides)** | | | |
| Mean (SD) | 0.063 (± 0.23) | 0.083 (± 0.26) | 0.077 (± 0.25) |
| Missing | 1 (1.4%) | 1 (0.7%) | 2 (0.9%) |
| **Diazole (Herbicides)** | | | |
| Mean (SD) | 0.014 (± 0.12) | 0.0036 (± 0.042) | 0.0071 (± 0.077) |
| **Triazine (Herbicides)** | | | |
| Mean (SD) | 0.32 (± 0.46) | 0.27 (± 0.44) | 0.29 (± 0.45) |
| Missing | 1 (1.4%) | 1 (0.7%) | 2 (0.9%) |
| **Triazinone (Herbicides)** | | | |
| Mean (SD) | 0 (± 0) | 0.014 (± 0.095) | 0.0095 (± 0.077) |
| Missing | 0 (0%) | 1 (0.7%) | 1 (0.5%) |
| **Traizole (Herbicides)** | | | |

**Table S1 – continued from previous page**

| | Case | Control | Total |
|---|---|---|---|
| | (N=72) | (N=139) | (N=211) |
| Mean (SD) | 0.061 (± 0.23) | 0.062 (± 0.23) | 0.061 (± 0.23) |
| Missing | 1 (1.4%) | 1 (0.7%) | 2 (0.9%) |
| **Amine (Herbicides)** | | | |
| Mean (SD) | 0.070 (± 0.24) | 0.095 (± 0.28) | 0.087 (± 0.27) |
| Missing | 1 (1.4%) | 0 (0%) | 1 (0.5%) |
| **Amide (Herbicides)** | | | |
| Mean (SD) | 0.099 (± 0.28) | 0.15 (± 0.34) | 0.13 (± 0.32) |
| Missing | 2 (2.8%) | 1 (0.7%) | 3 (1.4%) |
| **Urea or Sulfonylurea (Herbicides)** | | | |
| Mean (SD) | 0.18 (± 0.36) | 0.17 (± 0.36) | 0.18 (± 0.36) |
| Missing | 2 (2.8%) | 1 (0.7%) | 3 (1.4%) |
| **Quaternary ammonium (Herbicides)** | | | |
| Mean (SD) | 0.16 (± 0.36) | 0.15 (± 0.35) | 0.15 (± 0.35) |
| Missing | 2 (2.8%) | 0 (0%) | 2 (0.9%) |
| **Glyphosate (Herbicides)** | | | |
| Mean (SD) | 0.19 (± 0.38) | 0.14 (± 0.34) | 0.16 (± 0.36) |
| Missing | 2 (2.8%) | 0 (0%) | 2 (0.9%) |
| **Benzofuran (Herbicides)** | | | |
| Mean (SD) | 0.017 (± 0.12) | 0.019 (± 0.097) | 0.018 (± 0.10) |
| **Diphenyl ether (Herbicides)** | | | |
| Mean (SD) | 0 (± 0) | 0.027 (± 0.15) | 0.018 (± 0.12) |
| **Other (Herbicides)** | | | |
| Mean (SD) | 0.018 (± 0.12) | 0.014 (± 0.092) | 0.016 (± 0.10) |
| Missing | 1 (1.4%) | 0 (0%) | 1 (0.5%) |
| **DDT (Insecticides)** | | | |
| Mean (SD) | 0.11 (± 0.28) | 0.11 (± 0.28) | 0.11 (± 0.28) |
| Missing | 0 (0%) | 1 (0.7%) | 1 (0.5%) |
| **Lindane (Insecticides)** | | | |
| Mean (SD) | 0.22 (± 0.40) | 0.15 (± 0.33) | 0.17 (± 0.36) |
| Missing | 0 (0%) | 1 (0.7%) | 1 (0.5%) |
| **Dithiocarbamate (Fungicides)** | | | |
| Mean (SD) | 0.23 (± 0.40) | 0.16 (± 0.35) | 0.18 (± 0.37) |
| Missing | 1 (1.4%) | 1 (0.7%) | 2 (0.9%) |
| **Carbamate (Fungicides)** | | | |
| Mean (SD) | 0.16 (± 0.35) | 0.12 (± 0.31) | 0.13 (± 0.32) |
| Missing | 1 (1.4%) | 1 (0.7%) | 2 (0.9%) |
| **Dinitrophenol (Fungicides)** | | | |

**Table S1 – continued from previous page**

| | Case | Control | Total |
|---|---|---|---|
| | (N=72) | (N=139) | (N=211) |
| Mean (SD) | 0.021 (± 0.13) | 0.022 (± 0.15) | 0.021 (± 0.14) |
| **Aromatic (Fungicides)** | | | |
| Mean (SD) | 0.0028 (± 0.024) | 0.017 (± 0.12) | 0.012 (± 0.099) |
| Missing | 1 (1.4%) | 1 (0.7%) | 2 (0.9%) |
| **Triazole (Fungicides)** | | | |
| Mean (SD) | 0.092 (± 0.28) | 0.093 (± 0.28) | 0.093 (± 0.28) |
| Missing | 1 (1.4%) | 1 (0.7%) | 2 (0.9%) |
| **Dinitrophenol (Herbicides)** | | | |
| Mean (SD) | 0.24 (± 0.40) | 0.16 (± 0.35) | 0.19 (± 0.37) |
| Missing | 1 (1.4%) | 0 (0%) | 1 (0.5%) |
| **Diphenyl ether only (Herbicides)** | | | |
| Mean (SD) | 0.017 (± 0.12) | 0.013 (± 0.090) | 0.014 (± 0.10) |
| **Benzonitrile (Herbicides)** | | | |
| Mean (SD) | 0.066 (± 0.22) | 0.033 (± 0.15) | 0.044 (± 0.18) |
| Missing | 1 (1.4%) | 1 (0.7%) | 2 (0.9%) |
| **Aryloxyphenoxypropionic (Herbicides)** | | | |
| Mean (SD) | 0.038 (± 0.17) | 0.038 (± 0.17) | 0.038 (± 0.17) |
| Missing | 1 (1.4%) | 1 (0.7%) | 2 (0.9%) |
| **Urea (Herbicides)** | | | |
| Mean (SD) | 0.18 (± 0.36) | 0.17 (± 0.35) | 0.17 (± 0.35) |
| Missing | 2 (2.8%) | 1 (0.7%) | 3 (1.4%) |
| **Sulfonylurea (Herbicides)** | | | |
| Mean (SD) | 0.031 (± 0.17) | 0.030 (± 0.17) | 0.030 (± 0.17) |
| **Total carbamate** | | | |
| Mean (SD) | 0.30 (± 0.44) | 0.27 (± 0.43) | 0.28 (± 0.43) |
| Missing | 1 (1.4%) | 1 (0.7%) | 2 (0.9%) |
| **Total urea** | | | |
| Mean (SD) | 0.20 (± 0.37) | 0.17 (± 0.36) | 0.18 (± 0.36) |
| Missing | 1 (1.4%) | 1 (0.7%) | 2 (0.9%) |
| **Total amide** | | | |
| Mean (SD) | 0.17 (± 0.37) | 0.18 (± 0.37) | 0.18 (± 0.37) |
| Missing | 2 (2.8%) | 1 (0.7%) | 3 (1.4%) |
| **Total amine** | | | |
| Mean (SD) | 0.075 (± 0.24) | 0.096 (± 0.28) | 0.089 (± 0.27) |
| Missing | 1 (1.4%) | 0 (0%) | 1 (0.5%) |
| **Total triazine** | | | |
| Mean (SD) | 0.32 (± 0.46) | 0.28 (± 0.44) | 0.29 (± 0.45) |

**Table S1 – continued from previous page**

| | Case | Control | Total |
|---|---|---|---|
| | (N=72) | (N=139) | (N=211) |
| Missing | 1 (1.4%) | 1 (0.7%) | 2 (0.9%) |
| **Total diazine** | | | |
| Mean (SD) | 0.063 (± 0.23) | 0.083 (± 0.26) | 0.077 (± 0.25) |
| Missing | 1 (1.4%) | 1 (0.7%) | 2 (0.9%) |
| **Total phenol** | | | |
| Mean (SD) | 0.25 (± 0.41) | 0.18 (± 0.37) | 0.20 (± 0.38) |
| **Domestic use** | | | |
| Mean (SD) | 0.22 (± 0.42) | 0.19 (± 0.40) | 0.20 (± 0.40) |

**Table S2. Associations between sex and DNAm in TERRE samples accounting for $GRS_{pT=5e-8}$ and its interaction with sex.** $\Delta M$ is the $\log_2$-fold change in DNAm M-value observed. See Supplementary Data for all associations.

| Probe | CHR | POS | Gene | $\beta$ | t | $\Delta M$ | *p* value | Adjusted *p* value |
|---|---|---|---|---|---|---|---|---|
| cg11556740 | chr7 | 40586823 | *SUGCT* | 7.60E+00 | 5.80E+00 | 6.26E-01 | 2.48E-08 | 2.11E-06 |
| cg00968183 | chr17 | 66137457 | Intergenic | 3.40E+00 | 4.91E+00 | 4.11E-01 | 1.86E-06 | 7.92E-05 |
| cg21327887 | chr17 | 43971928 | *MAPT* | 2.89E+00 | 4.79E+00 | 3.05E-01 | 3.19E-06 | 9.03E-05 |
| cg20099416 | chr17 | 43971919 | *MAPT* | 2.42E+00 | 4.68E+00 | 2.87E-01 | 5.16E-06 | 1.10E-04 |
| cg18410271 | chr17 | 43472435 | *ARHGAP27* | 8.57E-01 | -4.30E+00 | -1.68E-01 | 2.63E-05 | 4.47E-04 |
| cg08582629 | chr18 | 21719467 | *CABYR* | 3.20E-01 | -4.16E+00 | -3.15E-01 | 4.61E-05 | 6.53E-04 |
| cg03727500 | chr2 | 232348334 | Intergenic | -1.15E-01 | -4.05E+00 | -3.63E-01 | 7.28E-05 | 8.85E-04 |
| cg16520312 | chr17 | 43971471 | *MAPT* | -2.69E-01 | -4.01E+00 | -3.33E-01 | 8.57E-05 | 9.10E-04 |
| cg20059597 | chr17 | 43726219 | *MGC57346-CRHR1* | -2.16E+00 | -3.47E+00 | -2.87E-01 | 6.36E-04 | 5.83E-03 |
| cg23202277 | chr17 | 43971911 | *MAPT* | -2.23E+00 | 3.45E+00 | 2.19E-01 | 6.85E-04 | 5.83E-03 |
| cg09764761 | chr17 | 44105544 | *MAPT* | -2.77E+00 | -3.28E+00 | -2.60E-01 | 1.22E-03 | 9.44E-03 |
| cg10780632 | chr17 | 43973522 | *MAPT* | -2.96E+00 | 3.22E+00 | 2.67E-01 | 1.51E-03 | 1.07E-02 |
| cg04226788 | chr17 | 44014390 | *MAPT* | -3.13E+00 | 3.16E+00 | 2.73E-01 | 1.81E-03 | 1.18E-02 |
| cg05727186 | chr17 | 43697356 | *MGC57346* | -3.46E+00 | 3.05E+00 | 2.16E-01 | 2.58E-03 | 1.57E-02 |
| cg25445714 | chr16 | 67152775 | *C16orf70* | -3.60E+00 | 3.00E+00 | 1.85E-01 | 3.01E-03 | 1.70E-02 |
| cg13438899 | chr17 | 44368573 | Intergenic | -3.66E+00 | -2.98E+00 | -2.24E-01 | 3.20E-03 | 1.70E-02 |
| cg07368061 | chr17 | 44090862 | *MAPT* | -3.80E+00 | 2.93E+00 | 2.40E-01 | 3.72E-03 | 1.86E-02 |
| cg11559198 | chr2 | 232348794 | Intergenic | -4.11E+00 | -2.82E+00 | -3.96E-01 | 5.25E-03 | 2.48E-02 |
| cg05314706 | chr17 | 43508250 | *ARHGAP27* | -4.29E+00 | -2.76E+00 | -2.09E-01 | 6.37E-03 | 2.71E-02 |
| cg05301556 | chr17 | 43971177 | *MAPT* | -4.27E+00 | -2.76E+00 | -2.53E-01 | 6.22E-03 | 2.71E-02 |
| cg16132033 | chr17 | 44292685 | *KANSL1* | -4.60E+00 | 2.64E+00 | 2.06E-01 | 8.97E-03 | 3.63E-02 |
| cg21214508 | chr17 | 44248233 | *KIAA1267* | -4.79E+00 | 2.56E+00 | 2.38E-01 | 1.12E-02 | 4.31E-02 |
